# Dynamics of COVID-19 pandemic in India and Pakistan: A metapopulation modelling approach

**DOI:** 10.1101/2021.08.02.21261459

**Authors:** Samantha J. Brozak, Binod Pant, Salman Safdar, Abba B. Gumel

## Abstract

India has been the latest global epicenter for COVID-19, a novel coronavirus disease that emerged in China in late 2019. We present a base mathematical model for the transmission dynamics of COVID-19 in India and its neighbour, Pakistan. The base model, which takes the form of a deterministic system of nonlinear differential equations, is parameterized using cumulative COVID-19 mortality data from each of the two countries. The model was used to assess the population-level impact of the control and mitigation strategies implemented in the two countries (notably community lockdowns, use of face masks, and social-distancing). Numerical simulations of the basic model indicate that, based on the current baseline levels of the control and mitigation strategies implemented, the pandemic trajectory in India is on a downward trend (as characterized by the reproduction number of the disease dynamics in India below, but close to, unity). This downward trend will be reversed, and India will be recording mild outbreaks (i.e., pandemic waves), if the control and mitigation strategies are relaxed from their current levels (e.g., relaxed to the extent that the associated community transmission parameters are increased by 20% or 40% from their current baseline values). Our simulations suggest that India could record up to 460,000 cumulative deaths by early September 2021 under the baseline levels of the control strategies implemented (up to 25,000 of the projected deaths could be averted if the control and mitigation measures are strengthened to the extent that the associated community transmission parameters are reduced by 20% from their baseline values). Our simulations show that the pandemic in Pakistan is much milder, with an estimated projected cumulative mortality of about 24,000 by early September 2021 under the baseline scenario. The basic model was extended to assess the impact of back-and-forth mobility between the two countries. Simulations of the resulting metapopulation model show that the burden of the COVID-19 pandemic in Pakistan increases with increasing values of the average time residents of India spend in Pakistan. In particular, it is shown that the India-to-Pakistan mobility pattern may trigger a fourth wave of the pandemic in Pakistan (under certain mobility scenarios), with daily mortality peaking in mid-August to mid-September of 2021. Under the respective baseline control scenarios, our simulations show that the back-and-forth mobility between India and Pakistan could delay the time-to-elimination of the COVID-19 pandemic in the two countries by three to five months (specifically, under the respective baseline scenarios, elimination could be delayed in India and Pakistan to November 2022 and July 2022, respectively).

## 1 Introduction

A new coronavirus disease emerged out of Wuhan, China, in December of 2019 [1, 2]. The disease, known as COVID-19 and caused by SARS-CoV-2, rapidly spread to every region of the world resulting in a global pandemic on a scale not seen since the 1918 influenza pandemic. As of June 14, 2021, COVID-19 accounted for 175.5 million confirmed cases and 3.4 million deaths globally [3]. India, one of the most populous countries in the world (with an estimated population of 1.3 billion people [4]), is currently the global epicenter for the disease. India reported its index case on January 27, 2020 [5], and, as of June 14, 2021, India recorded over 29.5 million confirmed cumulative cases and 375,000 COVID-19 deaths (India currently accounts for 95% of COVID-19 cases in South-East Asia) [6, 7]. India’s skyrocketing cumulative confirmed cases and COVID-19 mortality is associated with the onset of the second wave of the pandemic there in March of 2021, claiming over 183,000 lives thus far [7]. Neighbouring countries, such as Pakistan, have been on high-alert due to this second wave. As of June 14, 2021, Pakistan has recorded over 21,000 cumulative deaths due to COVID-19 since the onset of the pandemic began there in early 2020, and experienced a comparatively mild third wave in the spring of 2021 [7].

The symptoms of COVID-19 often resemble those of seasonal influenza, which typically include fever, difficulty breathing, fatigue, sore throat, and body aches [8]. The elderly and those with underlying medical conditions (comorbidities) are at higher risk for severe disease progression [9]. A significant percentage of individuals infected with COVID-19 experience mild or no clinical symptoms of the disease, and typically recover within two weeks [9]; some infected individuals suffer fatigue, headaches, shortness of breath, and other symptoms four weeks (or longer) after initial infection (known as “long COVID”) [10, 11]. Recent clinical studies have also highlighted the need for further research to investigate the effects of COVID-19 on the cardiovascular system [12, 13].

Prior to December 2020, when two vaccines were given Emergency Use Authorization (EUA) by the Food and Drugs Administration (FDA) of the United States [14, 15], control measures against COVID-19 were hitherto focused on the use of nonpharmaceutical interventions (NPIs), such as quarantine of suspected cases, isolation of people with clinical symptoms of the disease, community lockdowns, the use of face masks (or face coverings) in public, physical-distancing, etc. [16–19]. A number of antivirals and monoclonal antibody therapies were also developed and used to treat cases of severe illness; however, *remdesivir* is currently the only FDA-approved antiviral for treating COVID-19 [20–23].

In jurisdictions with low vaccination coverages, the use of NPIs has been emphasized to reduce community transmission and burden of COVID-19 [16, 19, 24]. For instance, several regions in India and Pakistan have intensified the implementation of NPIs, including lockdowns, during the ongoing second wave of the pandemic [25, 26]. Specifically, some local authorities (or local governments) banned large social gatherings, closed certain businesses and schools, in addition to implementing curfews and strongly encouraging the use of face masks in public, in an effort to mitigate the burden of the second wave of the pandemic [25, 26]. Although a nationwide lockdown was not enforced in India, several of the most populous states (such as Maharashtra and Uttar Pradesh) implemented lockdowns due to the rising number of confirmed cases during mid- to late April of 2021 (with some states easing restrictions in June, 2021) [26–30]. Pakistan, on the other hand, announced a ten-day nationwide lockdown beginning May 8, 2021, with some of the hardest-hit cities and districts announcing restrictions from late March to mid April [25, 31–33]. On April 21, 2021, the military was deployed in several cities in Pakistan to ensure that precautionary measures against COVID-19 were followed [34]. Several states in Pakistan began reopening schools in early June due to a decline in cases [35].

The second wave of the pandemic in India, which has caused such unprecedented devastation (and severely overwhelmed the nation’s healthcare system) [36, 37], was due to the emergence of a new COVID-19 variant (known as the Delta B.1.617 variant), which is more readily transmissible and virulent than the original SARS-CoV-2 strain that hit India during the first wave of the pandemic [38, 39]. Specifically, the increased prevalence of the B.1.617 variant (which was first detected in India in February 2021; *albeit* its sub-lineages (B.1.617.1, B.1.617.2, and B.1.617.3) were detected as far back as October 2020 [38, 39]) and its sub-lineages coincide with a surge of cases in India [38]. Sub-lineages of the B.1.617 variant appear to have higher rates of transmission and reduced antibody neutralization [38, 39], causing the B.1.617 variant to be declared a *a variant of concern* by the World Health Organization and *a variant of interest* by the Centers for Disease Control and Prevention of the United States [39].

Mathematical models of various types have been developed and used to study the transmission dynamics of COVID-19, and to assess various control and mitigation strategies [18, 19, 40–44]. For instance, Li *et al*. [40] used a metapopulation model to estimate the proportion of asymptomatic or mild infections and determine the relative infectiousness of these undocumented cases. They reported that approximately 86% of cases were unaccounted for prior to the implementation of travel restrictions, and these undocumented cases were significant drivers of documented cases. After mitigation efforts were put in place (isolation and travel restrictions), more than half of cases were accounted for [40]. Similarly, Verity *et al*. [41] used a statistical model to deduce that nearly half of cases in China were unidentified in the early stages of the pandemic. Results from a stochastic model developed by Kucharski *et al*. [42] showed that travel restrictions can reduce transmission and prevent outbreaks.

Ngonghala *et al*. [18] developed a comprehensive deterministic model for assessing the population-level impacts of nonpharmaceutical interventions, such as community lockdowns, masking, and various testing strategies. Their study showed that even moderate face mask adoption could prevent a resurgence of COVID-19 post-lockdown and highlighted the importance of detecting asymptomatic infections. Similarly, a model developed by Iboi *et al*. [43] to capture disease dynamics in Nigeria indicated that premature relaxation of lockdown measures may cause a second wave. Eikenberry *et al*. [19] found that moderately effective masks (such as cloth face coverings) can significantly reduce COVID-19 transmission and subsequent mortality.

Globalization and the importance of travel restrictions found in previous work highlight the need to study COVID-19 transmission in an increasingly-connected world. Lee *et al*. [45] developed a two-patch SIR model (susceptible-infected-recovered) with travel defined using a residence-time matrix. Their study indicated that controlling epidemics in both patches at the same time is the most effective strategy to reduce the final size of the epidemic, and that results are dependent on the residence-time matrix and transmission rates. Bichara *et al*. [46] found similar dependence of the final epidemic size on the residence-time matrix.

The devastating impact of B.1.617 COVID-19 variant, owing to its increased transmissibility and reduced antibody neutralization (in comparison to the previous strain that was circulating in India), has motivated the urgent need to use mathematical modeling, coupled with statistical data analytics and computation, to assess the public health impact and burden of this COVID-19 variant, and to devise effective control and mitigation strategies. Further, it is imperative to assess the potential impact of the disease dynamics in India on its neighbouring countries (particularly Pakistan), which are potentially quite vulnerable to recording higher COVID-19 burden due to their proximity to the current global epicenter. Consequently, the objective of the current study is to use mathematical modeling approaches to study the transmission dynamics of the COVID-19 pandemic, and evaluate the impact of control and mitigation measures, in both India and Pakistan, and assess the potential impact of the back-and-forth mobility of individuals between the two countries. In particular, a basic Kermack-McKendrick-type epidemic model will be developed and used to study the dynamics in each of the two countries. The basic model will be extended to a metapopulation model, with Lagrangian residence time, to account for the impact of the back-and-forth mobility between the two countries. The paper is organized as follows. The basic model is formulated in Section 2. Its basic qualitative properties, as well as its fitting to observed cumulative mortality data and the asymptotic stability analyses of its disease-free equilibria, are also reported. The metapopulation model is formulated in Section 3.

## 2 Formulation of a Basic Model for COVID-19 Dynamics

The basic epidemic model for the transmission-dynamics of COVID-19 in a population is developed by stratifying the total population at time *t*, denoted by *N* (*t*), into the mutually-exclusive compartments of susceptible individuals (*S*(*t*)), exposed or latent individuals (i.e., newly-infected individuals who are not yet infectious; *E*(*t*)), pre-symptomatic infectious individuals (*P* (*t*)), symptomatically-infectious individuals (*I*(*t*)), asymptomatically-infectious individuals (*A*(*t*)), hospitalized individuals (*H*(*t*)), and recovered individuals (*R*(*t*)), so that *N* (*t*) = *S*(*t*) + *E*(*t*) + *P* (*t*) + *I*(*t*) + *A*(*t*) + *H*(*t*) + *R*(*t*). Individuals in the exposed (or latent) compartment, *E*(*t*), are those who are newly-infected with the disease but are not yet able to transmit the disease (i.e., they are not yet infectious). The basic model is given by the following deterministic system of nonlinear differential equations (where a dot represents differentiation with respect to time *t*):

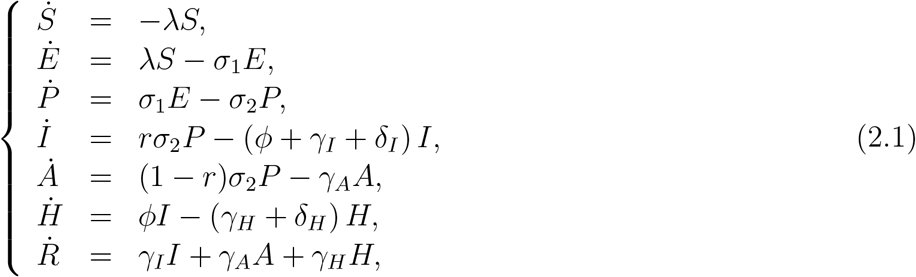

where *λ* is the *force of infection* and is defined as 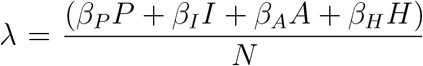, with *β*_*P*_, *β*_*I*_, *β*_*A*_ and *β*_*H*_ representing, respectively, the effective contact rate for individuals in the *P, I, A* and *H* compartment (where *N* is the total population size).

In the basic model (2.1), susceptible individuals acquire COVID-19 infection, following effective contacts with individuals in the *P, I, A* and *H* classes, at the rate *λ*. The parameter *σ*_1_ represents the progression rate of individuals in the exposed (*E*) class to the pre-symptomatically-infectious class (*P*). Similarly, *σ*_2_ represents the progression rate of infectious individuals in the *P* class to either the asymptomatically-infectious class (*A*) or to the symptomatically-infectious class (*I*). Specifically, a proportion, *r*, of these individuals progress to the symptomatically-infectious class (at a rate *rσ*_2_) and the remaining proportion, 1 *− r*, progress to the asymptomatically-infectious class (at a rate (1 *− r*)*σ*_2_). Therefore, the intrinsic incubation period of the disease is 1*/σ*_1_ + 1*/σ*_2_. The parameter *γ*_*I*_(*γ*_*A*_)(*γ*_*H*_) represents the recovery rate for infectious individuals in the *I*(*A*)(*H*) class. Furthermore, *ϕ* is the hospitalization rate of individuals with clinical symptoms of the disease. Finally, the parameter *δ*_*I*_(*δ*_*H*_) represents the disease-induced mortality rate for individuals in the *I*(*H*) class. We assume *β*_*P*_ ≠ *β*_*I*_ ≠ *β*_*A*_ ≠*β*_*H*_, which allows us to account for possible heterogeneity in the contact rates of infectious individuals in the pre-symptomatic (*P*), symptomatic (*I*), asymptomatic (*A*) and the hospitalized (*H*) classes. It should be noted that, although NPIs, such as masking and social-distancing are not explicitly accounted for in the basic model (2.1), these measures are embedded in the community contact rates (*β*_*P*_, *β*_*I*_, *β*_*A*_ and *β*_*H*_).

Figure 1 depicts the flow diagram of the basic model (2.1). The state variables and parameters of the basic model are described in Table 1 and Table 2, respectively. Some of the main assumptions made in the formulation of the basic model (2.1) include:

**Table 1:** Description of the state variables of the basic model (2.1).

| State variable | Description |
| --- | --- |
| $S$ | Number of susceptible individuals |
| $E$ | Number of exposed (newly-infected but not yet infectious) individuals |
| $P$ | Number of pre-symptomatically-infectious individuals |
| $I$ | Number of symptomatically-infectious individuals |
| $A$ | Number of asymptomatically-infectious individuals |
| $H$ | Number of hospitalized individuals |
| $R$ | Number of recovered individuals |

**Table 2:** Description of the parameters of the basic model (2.1).

| Parameter | Description |
| --- | --- |
| $\beta_P(\beta_I)(\beta_A)(\beta_H)$ | Effective contact rate of individuals in the $P(I)(A)(H)$ compartment |
| $\sigma_1$ | Progression rate from exposed to pre-symptomatic class (latent period) |
| $r$ | Proportion of individuals who show clinical symptoms of the disease |
| $r\sigma_2$ | Progression rate from the pre-symptomatic to the symptomatic infectious class |
| $(1-r)\sigma_2$ | Progression rate from the pre-symptomatic to the asymptomatic infectious class |
| $\phi$ | Hospitalization rate for the symptomatically-infectious individuals |
| $\gamma_I(\gamma_A)(\gamma_H)$ | Recovery rate for individuals in the $I(A)(H)$ class |
| $\delta_I(\delta_H)$ | Disease-induced mortality rate for individuals in the $I(H)$ compartment |

**Figure 1:**
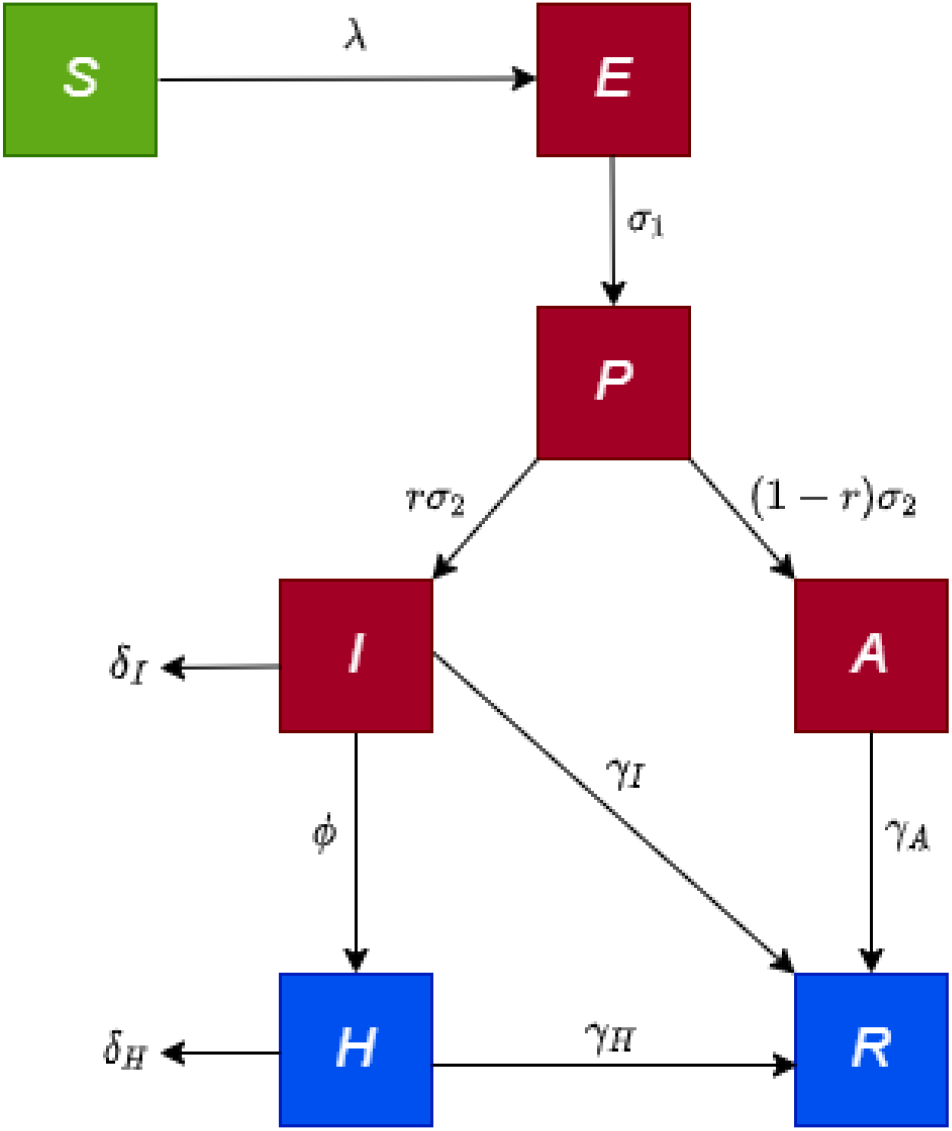
Flow diagram of the basic model (2.1).

a. a well-mixed, closed population (i.e., individuals are indistinguishable, and every member of the population can mix with every other member of the population);
b. exponentially-distributed waiting times in each epidemiological compartment;
c. the timescale of the novel pandemic is much shorter than the demographic timescale (so that births and natural death processes can be ignored);
d. natural recovery induces permanent immunity against future infection. Although there is currently limited evidence regarding the duration of infection-acquired immunity, Dan *et al*. [47] recently estimated that natural immunity due to recovery from COVID-19 could last up to eight months.

For housekeeping purposes, it is convenient to let *D*(*t*) be the total number of individuals who died from COVID-19 in the population. It follows from the basic model (2.1) that the equation for the rate of change of *D*(*t*) is 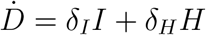.

The basic qualitative properties of the single-patch model (2.1) will now be explored.

### 2.1 Qualitative Properties of the Basic Model

Before carrying out asymptotic analysis and numerical simulations of the basic model (2.1), it is instructive that we explore the basic qualitative properties of the model (2.1). We define the following biologically-feasible region for the basic model (2.1):

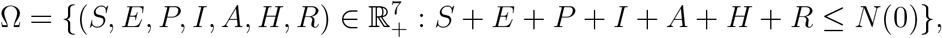

where *N* (0) is the initial total population. For the basic model (2.1) to be mathematically- and biologically-meaningful, it is necessary that the solutions of the basic model (2.1) remain non-negative for all non-negative initial conditions. That is, solutions that start in Ω remain in Ω for all time *t >* 0 (i.e., Ω is positively-invariant with respect to the basic model (2.1)) [17]. We claim the following result.

#### Theorem 2.1.

*Let S*(0) *>* 0, *E*(0) *≥* 0, *P* (0) *≥* 0, *I*(0) *≥* 0, *A*(0) *≥* 0, *H*(0) *≥* 0 *and R*(0) *≥* 0. *Then, S*(*t*) *>* 0, *E*(*t*) *≥* 0, *P* (*t*) *≥* 0, *I*(*t*) *≥* 0, *A*(*t*) *≥* 0, *H*(*t*) *≥* 0 *and R*(*t*) *≥* 0 *for all t >* 0.

The proof of Theorem 2.1 is given in Appendix A. Mathematically, Theorem 2.1 shows that the basic model (2.1) is well-posed in Ω. Hence, it is sufficient to study its dynamics in Ω.

### 2.2 Asymptotic Stability Analysis of Disease-free Equilibria

The basic model (2.1) has a continuum of disease-free equilibria (DFE), given by:

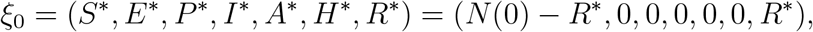

where *N* (0) is the initial total population, 0 *< S*^*∗*^ *≤ N* (0), 0 *≤ R*^*∗*^ *< N* (0), and 0 *< S*^*∗*^ + *R*^*∗*^ *≤ N* (0). The asymptotic stability property of *ξ*_0_ can be explored using the *next generation operator method* [48]. Using the notation in [48], it follows that the associated non-negative matrix of new infection terms (*F*) and the M-matrix of all linear transition terms (*V*) are given, respectively, by

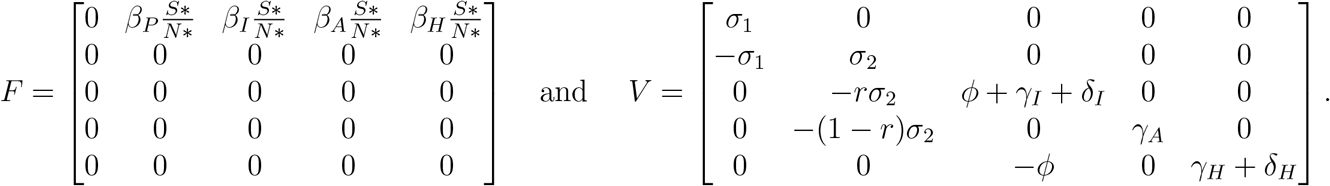

It is convenient to define the quantity (where *ρ* is the spectral radius):

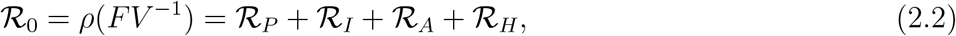

where,

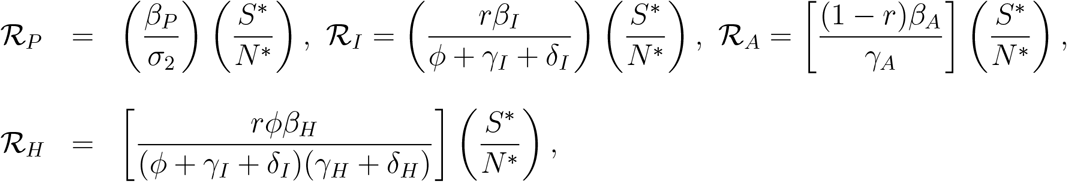

and *N* ^*∗*^ = *S*^*∗*^ + *R*^*∗*^ (with *S*^*∗*^ and *R*^*∗*^ as defined above). The quantity ℛ_0_ is the *basic reproduction number* of the basic model (2.1). It measures the average number of new cases generated by a typical infected individual if introduced into a completely susceptible population (i.e., a population where no one has immunity due to previous exposure to the disease and no public health intervention and mitigation measures are implemented in the community). The quantities ℛ_*P*_, ℛ_*I*_, ℛ_*A*_ and ℛ_*H*_ are the constituent basic reproduction numbers for the pre-symptomatic, symptomatic, asymptomatic, and hospitalized individuals, respectively. The result below follows from Theorem 2 of [48].

#### Theorem 2.2.

*The continuum of disease-free equilibria* (*ξ*_0_) *of the basic model* (2.1) *is locally-asymptotically stable (LAS) if ℛ*_0_ *<* 1.

Theorem 2.2 implies that a small influx of COVID-19 cases will not cause a significant outbreak in the community if *ℛ*_0_ *<* 1. In the case of epidemic models, such as the basic model (2.1), the epidemiological requirement of having ℛ_0_ *<* 1, while sufficient, is not necessary for the elimination of the disease from the community. This is owing to the fact that Kermack-McKendrick-type epidemic models (such as (2.1)) do not include vital dynamics (i.e., no birth and natural death processes) and no immigration (since the model assumes a closed population). In such models, the population of susceptible individuals is always decreasing (since there is no influx of new susceptible individuals into the community, by birth or immigration). Furthermore, in the formulation of the basic model (2.1), it was assumed that recovered individuals are immune from acquiring future infections (hence, in this setting, the virus will eventually have no susceptible host to infect). Consequently, the disease ultimately dies out even if the basic reproduction number (*ℛ*_0_) exceeds unity.

#### Remark

*In the case that ℛ*_0_ *>* 1, *the epidemic will grow to a peak and then eventually decline to zero [43, 44]*. *Furthermore, following [17], the ratio* 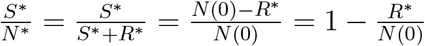 *in the expression for ℛ*_0_ *can be expressed as* 1*−f*_*r*_, *where* 0 *≤ f*_*r*_ *≤* 1 *is the proportion of the individuals who recovered from COVID-19 and have immunity against the disease [17]*. *Hence, as noted by [17], ℛ*_0_ *decreases with increasing values of f*_*r*_, *and ℛ*_0_ *→* 0 *as f*_*r*_ *→* 1 *(where* 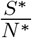 *is replaced by* 1 *− f*_*r*_ *in* (2.2)*), so that (epidemiologically) pandemic elimination is more likely as the proportion of recovered (and, thus, immune) individuals increases*.

The basic reproduction number of the basic model (2.1) is epidemiologically interpreted below.

#### 2.2.1 Epidemiological interpretation of the basic reproduction number

The quantity *ℛ*_*P*_ in the expression for *ℛ*_0_ is the product of the infection rate by pre-symptomatically infectious individuals near the continuum of disease-free equilibria (*β*_*P*_) and the average duration in the pre-symptomatically infectious class 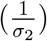. Similarly, *ℛ*_*I*_ is the product of the infection rate of symptomatically-infectious individuals near the disease-free equilibria (*β*_*I*_), the probability of surviving the pre-symptomatic class and moving to the symptomatic compartment 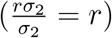 and the average duration in the symptomatically-infectious class 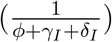. The quantity *ℛ*_*A*_ is the product of the infection rate of asymptomatically-infectious individuals (*β*_*A*_), the probability of surviving the pre-symptomatic class and moving to the asymptomatically-infectious class 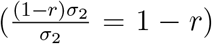 and the average time spent in the asymptomatically-infectious class 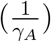. Finally, *ℛ*_*H*_ is the product of the transmission rate of hospitalized individuals (*β*_*H*_), the probability of surviving the pre-symptomatic infectious class and moving to the symptomatic infectious class 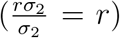, the probability of surviving the symptomatic class and moving to the hospitalized class 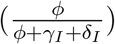 and the average duration in the hospitalized class 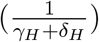.

The sum of *ℛ*_*P*_, *ℛ*_*I*_, *ℛ*_*A*_ and *ℛ*_*H*_ gives *ℛ*_0_. When referring to the reproduction number *ℛ*_0_ obtained via the basic model (2.1) in India and Pakistan, we use the notation *ℛ*_0*i*_ and *ℛ*_0*p*_, respectively.

In order to ensure that elimination of the pandemic when ℛ_0_ is less than unity is independent of the initial size of the sub-populations of the basic model (2.1), it is necessary to show that the continuum of disease-free equilibria (*ξ*_0_) is globally-asymptotically stable. We claim the following result:

##### Theorem 2.3.

*The continuum of the disease-free equilibria* (*ξ*_0_) *of the basic model* (2.1) *is globally-asymptotically stable (GAS) in* Ω *whenever ℛ*_0_ *≤* 1.

The proof of Theorem 2.3, based on using Lyapunov function theory and LaSalle’s Invariance Principle [49], is given in Appendix B. Epidemiologically-speaking, Theorem 2.3 shows that COVID-19 elimination is independent of the initial number of infected individuals introduced into the community.

### 2.3 Data-fitting and Parameter Estimation

Since the objective of this study is to analyse the transmission dynamics and control of COVID-19 in India and Pakistan, it is necessary to estimate the unknown parameters of the basic model (2.1) when used to simulate the disease dynamics in both India and Pakistan. To achieve this, we now fit the basic model with the cumulative COVID-19 mortality data for both India and Pakistan. Specifically, it should be recalled that the basic model (2.1) contains 13 parameters, the values of six of which (*σ*_1_, *r, σ*_2_, *γ*_*I*_, *γ*_*A*_ and *γ*_*H*_) are known from the literature (as tabulated in Table 3). The values of the remaining parameters (namely the contact rates, *β*_*P*_, *β*_*I*_, *β*_*A*_, and *β*_*H*_, the hospitalization rate, *ϕ*, and the disease-induced death rates, *δ*_*I*_ and *δ*_*H*_) are obtained from fitting the model with the cumulative mortality data for India and Pakistan, obtained from the Johns Hopkins University COVID-19 repository [7].

**Table 3:** Table of fixed parameters of the basic model (2.1) for India and Pakistan.

| Parameter | Value | Source |
| --- | --- | --- |
| $\sigma_1$ | $1/4 \text{ day}^{-1}$ | [44, 50] |
| $\sigma_2$ | $1/2 \text{ day}^{-1}$ | [44, 50] |
| $r$ | 0.6 (dimensionless) | [44, 50] |
| $1 - r$ | 0.4 (dimensionless) | [44, 50] |
| $\gamma_I$ | $1/10 \text{ day}^{-1}$ | [24] |
| $\gamma_A$ | $1/5 \text{ day}^{-1}$ | [51] |
| $\gamma_H$ | $1/14 \text{ day}^{-1}$ | [52, 53] |

#### 2.3.1 Fixed parameters

As stated above, the values of some of the parameters of the model (2.1) are obtained from the literature (and are tabulated in Table 3). Some of these parameter values are justified below. The overall incubation period of the disease is given by 1*/σ*_1_ + 1*/σ*_2_ = 6 days, with infected individuals able to transmit in the last 2 days of the incubation period (i.e., they are able to transmit during the presymptomatic infectious period) [44, 50]. Furthermore, the parameter *r* is given the value *r* = 0.6 to signify the fact that 60% of infected individuals will show clinical symptoms of COVID-19 (while the remaining 40% will not) [44, 50]. Symptomatically-infectious individuals recover at a rate of *γ*_*I*_ = 1*/*10 *per* day [24], while asymptotically-infectious individuals recover at a rate *γ*_*A*_ = 1*/*5 *per* day [51] and hospitalized individuals recover at a rate of *γ*_*H*_ = 1*/*14 *per* day [52, 53].

#### 2.3.2 Fitted (estimated) parameters for India and Pakistan

The basic model (2.1) will now be fitted to the cumulative mortality data for India and Pakistan, obtained from the Johns Hopkins University COVID-19 repository [7], for the period from March 1, 2021 to June 6, 2021. These dates are chosen because they coincide with the beginning of the “second wave” in India (which started in March 2021) until early June 2021. Specifically, MATLAB’s fmincon optimizer will be used to obtain the best values of the unknown parameters of the basic model (2.1) for each of the two countries. Sum squared error (SSE) is used to evaluate the goodness of fit [54]. It should be recalled that numerous jurisdictions in both India and Pakistan implemented lockdown measures (curfews, closing of businesses, etc. [25, 26]) in the spring of 2021, with start dates ranging from late March to mid-April [30, 35]. Consequently, we fitted our model using data for the pre-lockdown (estimated to be from March 1, 2021 to April 22, 2021) and lockdown (April 22, 2021 to June 6, 2021) periods for each country. Although numerous jurisdictions in India and Pakistan started lockdown at different times during the spring of 2021, we estimated the overall time for the onset of lockdown to be April 22, 2021 for each of the two nations.

The results of the fitting of the basic model (2.1) with the cumulative COVID-19 mortality for each country are depicted in Figure 2(a) for India and Figure 2(c) for Pakistan. Furthermore, the estimated values of the unknown parameters for India and Pakistan, obtained from the fitting of the basic model (2.1) with the respective cumulative mortality for each country, are tabulated in Table 4a (for India) and Table 4b (for Pakistan), showing fits that capture both cumulative and daily mortality. The plots depicted in Figure 2(a) (for India) and Figure 2(c) (for Pakistan) compare the model output to the cumulative COVID-19 mortality data, while Figure 2(b) and Figure 2(d) show the seven-day rolling average of the daily mortality generated by simulating the basic model with the fitted and fixed parameters for India and Pakistan, respectively. Using the parameter values listed in Table 3 and Table 4a, we find that the reproduction number for India, *ℛ*_0*i*_, is approximately 1.84 from March 1 through April 22, 2021, and reduces to *ℛ*_0*i*_ *≈* 0.79 from April 22 to June 6, 2021. Similarly, using the parameter values in Table 4b for Pakistan, ℛ_0*p*_ was estimated to be 1.46 from March 1 through April 22, 2021 and decreased to ℛ_0*p*_ *≈* 0.80 from April 22 to June 6, 2021. Thus, our estimates for the reproduction numbers of each country, before and during lockdown, indicate that the implementation of lockdown measures were successful in effectively curtailing the size of outbreaks in each country (by bringing the respective reproduction number below one). It is also worth noting that the value of the reproduction number for the COVID-19 epidemic in India (ℛ_0*i*_), before lockdown is higher than that for Pakistan (ℛ_0*p*_) and even during the lockdown it is approximately very close to the reproduction number of Pakistan. Hence, India has consistently been recording larger (and more severe) COVID-19 outbreaks, in comparison to its neighbor, Pakistan.

**Table 4:**
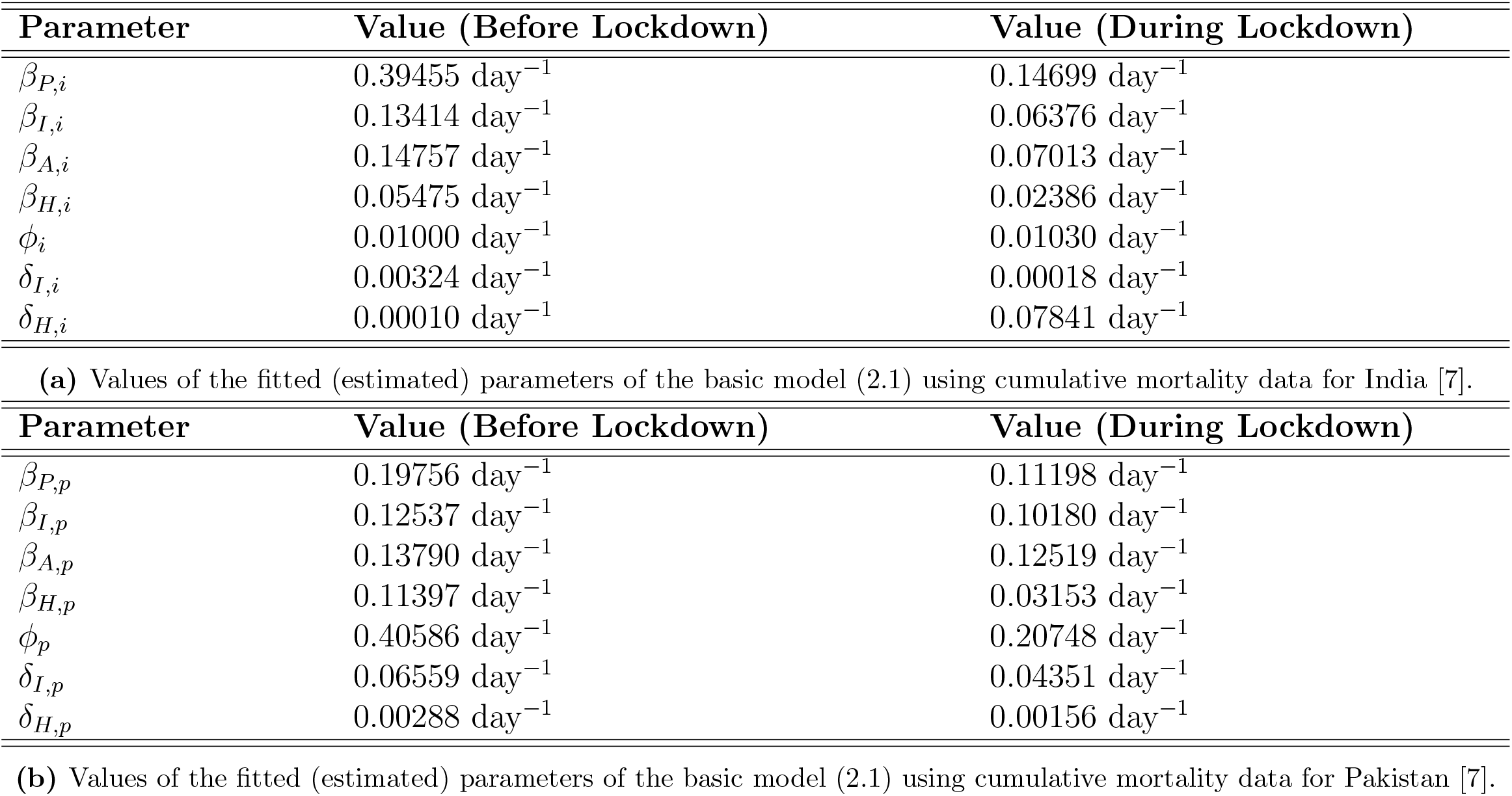
Values of fitted (estimated) parameters of the basic model (2.1) for (a) India and (b) Pakistan, obtained using cumulative mortality data from the Johns Hopkins University COVID-19 Repository [7]. Subscript *i* denotes a parameter estimated using the cumulative mortality data for India, and subscript *p* denotes a parameter estimated using the cumulative mortality data for Pakistan.

**Figure 2:**
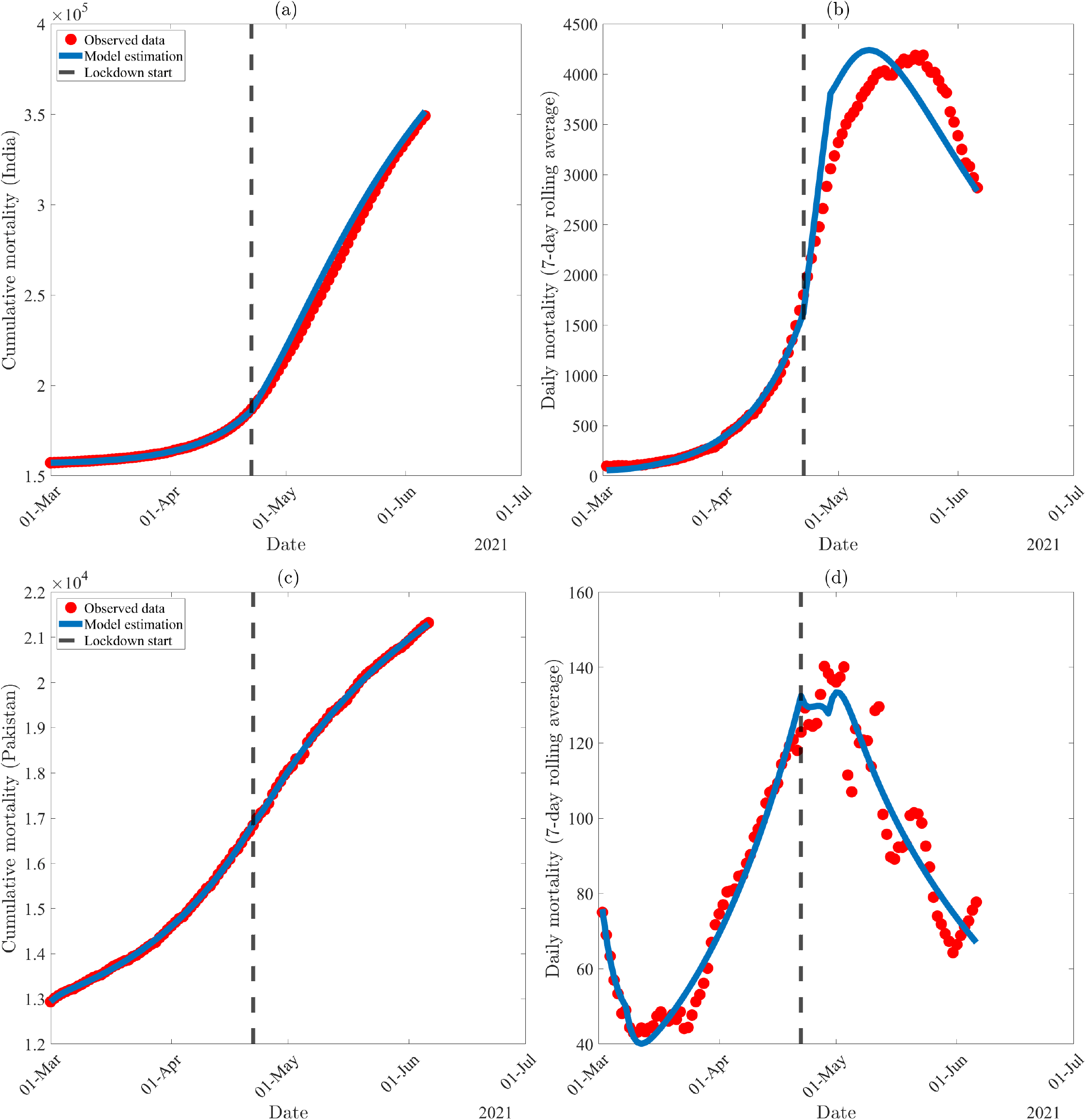
Data-fitting results of the basic model (2.1), showing data and model output for (a) cumulative COVID-19-induced mortality in India, (b) the seven-day rolling average for daily mortality in India, (c) cumulative COVID-19-induced mortality in Pakistan, and (d) the seven-day rolling average for daily mortality in Pakistan. Red dots indicate data points, while the solid line represents output from the model (2.1) for the period from March 1, 2021 through June 6, 2021. The dashed line represents the date for onset of lockdown (April 22, 2021). Values for fitted parameters are listed in Table 4.

### 2.4 Numerical Simulations of the Basic Model

Although the lockdown measures were implemented in the two countries around April 22, 2021, numerous regions within the two countries have begun relaxing these measures by early June 2021 [30, 35]. It is therefore instructive to assess the potential impact of such strengthening and relaxation of the lockdown measures on the trajectory of the pandemic in the two nations. To achieve this objective, the basic model (2.1) will be simulated, unless otherwise stated, using the baseline values of the fixed and fitted parameters for India (tabulated in Tables 3 and 4a) and Pakistan (given in Tables 3 and 4b). The simulations are aimed at assessing the population-level impact of relaxation and strengthening of the lockdown measures in the two countries. Specifically, we will use the baseline scenario (i.e., we will use baseline values of each parameter of the model, based on the aforementioned fixed parameters and fitted parameters obtained from fitting the basic model with the COVID-19 mortality data for the period March 1, 2021 to June 6, 2021) to predict the trajectory of the COVID-19 pandemic in each of the two countries until September 4, 2021. The impact of the relaxation or strengthening of the lockdown measures is measured by decreasing or increasing the baseline values of the community contact rate parameters of the basic model (2.1), namely *β*_*P*_, *β*_*I*_, *β*_*A*_, and *β*_*H*_, by a certain percentage.

Figure 3(a) depicts the cumulative COVID-19 mortality for India, as a function of time, for various effectiveness levels of the lockdown measures implemented in India during the second wave of the pandemic. This figure shows that, under the baseline scenario (i.e., if the baseline values of the fitted and estimated parameters from the lockdown period are used in the simulations), India will record a projected 459, 000 cumulative deaths by September 4, 2021 (Figure 3(a), gold curve). For the baseline scenario, the associated basic reproduction number for the dynamics of the disease in India is estimated to be *ℛ*_0*i*_ *≈* 0.79 (suggesting that, under the baseline scenario, the pandemic is on a downward trajectory in India, since *ℛ*_0*i*_ *<* 1). This figure further shows that the projected cumulative mortality decreases with increasing effectiveness level of the lockdown measures. For instance, if the lockdown measures in India are strengthened to the extent that the community contact rate parameters are decreased by 20%, our simulations (Figure 3(a), purple curve) show that up to 25, 000 of the projected 459, 000 deaths by September 4, 2021 can be averted (this represents approximately a 5.4% decrease in the projected cumulative mortality). Under this scenario, the basic reproduction number decreases from the baseline value of 0.79 to ℛ_0*i*_ *≈* 0.63 (thereby accelerating the prospect of pandemic elimination in India). Thus, our simulations show that increasing the effectiveness levels of the lockdown measures in India by 20% (from the current baseline level) could have averted tens of thousands of deaths, in addition to accelerating pandemic elimination prospect in India.

**Figure 3:**
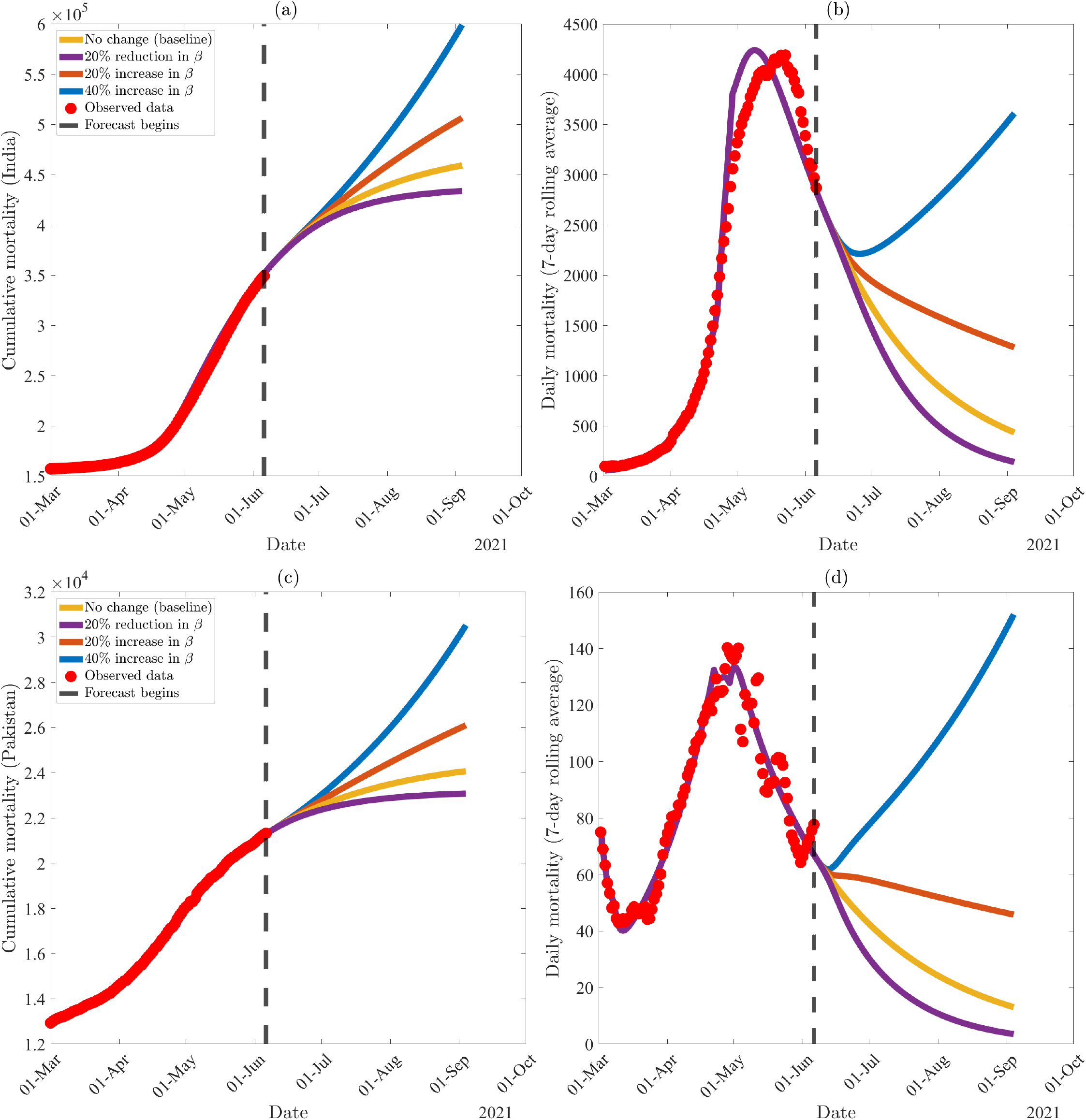
Numerical simulations of the basic model (2.1) for various effectiveness levels of the implementation or relaxation of community lockdown measures, showing (a) projected cumulative mortality for India, (b) the seven-day rolling average of daily mortality in India, (c) projected cumulative mortality for Pakistan, and (d) the seven-day rolling average of daily mortality for Pakistan. Values for fixed parameters are listed in Table 3. Values of the estimated parameters prior to June 6, 2021 are listed in Table 4a (India) and Table 4b (Pakistan). Values for the estimated parameters after June 6, 2021 are computed by increasing or decreasing the baseline values of the estimated contact rates (*β*_*P*_, *β*_*I*_, *β*_*A*_, *β*_*H*_) by the indicated percentage. Values estimated during the lockdown period are used for other estimated parameters (*ϕ, δ*_*I*_, *δ*_*H*_). Red dots represent cumulative mortality data for the respective countries [7], and the dashed solid line represents the date where 90-day forecasts begin.

On the other hand, relaxing the lockdown measures (from their current baseline levels) will result in a significant increase in the projected cumulative mortality. For instance, if the relaxation of lockdown measures resulted in a 20% increase in the baseline values of the community contact rate parameters, the projected cumulative mortality for India on September 4, 2021 will increase by 48, 000 (Figure 3(a), brown curve), representing a 10.5% increase in the projected baseline cumulative mortality (this corresponds to an increase in ℛ_0*i*_ from 0.79 at the baseline to ℛ_0*i*_ *≈* 0.95; thereby increasing the likelihood of more severe outbreaks). If the relaxation of lockdown measures is further increased to correspond to a 40% increase in the baseline values of the community contact rate parameters, our simulations show that up to 140, 000 additional deaths could be recorded in India by September 4, 2021 (Figure 3(a), blue curve), representing approximately a 30.5% increase in the projected cumulative mortality. Under this scenario, the reproduction number increases to *ℛ*_0*i*_ *≈* 1.11 (suggesting that India could experience a third wave of the pandemic, which would be relatively mild since ℛ_0*i*_ is only slightly above one). In other words, the simulations in Figure 3 show that relaxing the effectiveness levels of the lockdown measures implemented in India, from their current baseline level to a level that can cause a 20% or 40% increase in the baseline values of the community contact rate parameters, would cause a dramatic increase in the projected cumulative mortality (i.e. 48, 000 deaths and 140, 000 deaths, respectively) to be recorded in India by September 4, 2021, in addition to increasing the likelihood of a third wave of the pandemic in India. Thus, these simulations suggest that relaxing the current (baseline) level of the lockdown measures implemented in India, to the aforementioned levels (particularly the case where relaxation of lockdown corresponds to 40% increase in baseline levels of community contact rate parameters), could trigger a relatively mild third wave of the pandemic in India. For this scenario, where lockdown measures are relaxed to correspond to a 40% increase in the community contact rates, our simulations show that the predicted third wave of the pandemic in India could peak by June 2022 (Figure 4(a)), while the daily mortality could peak a month later (Figure 4(b)).

**Figure 4:**
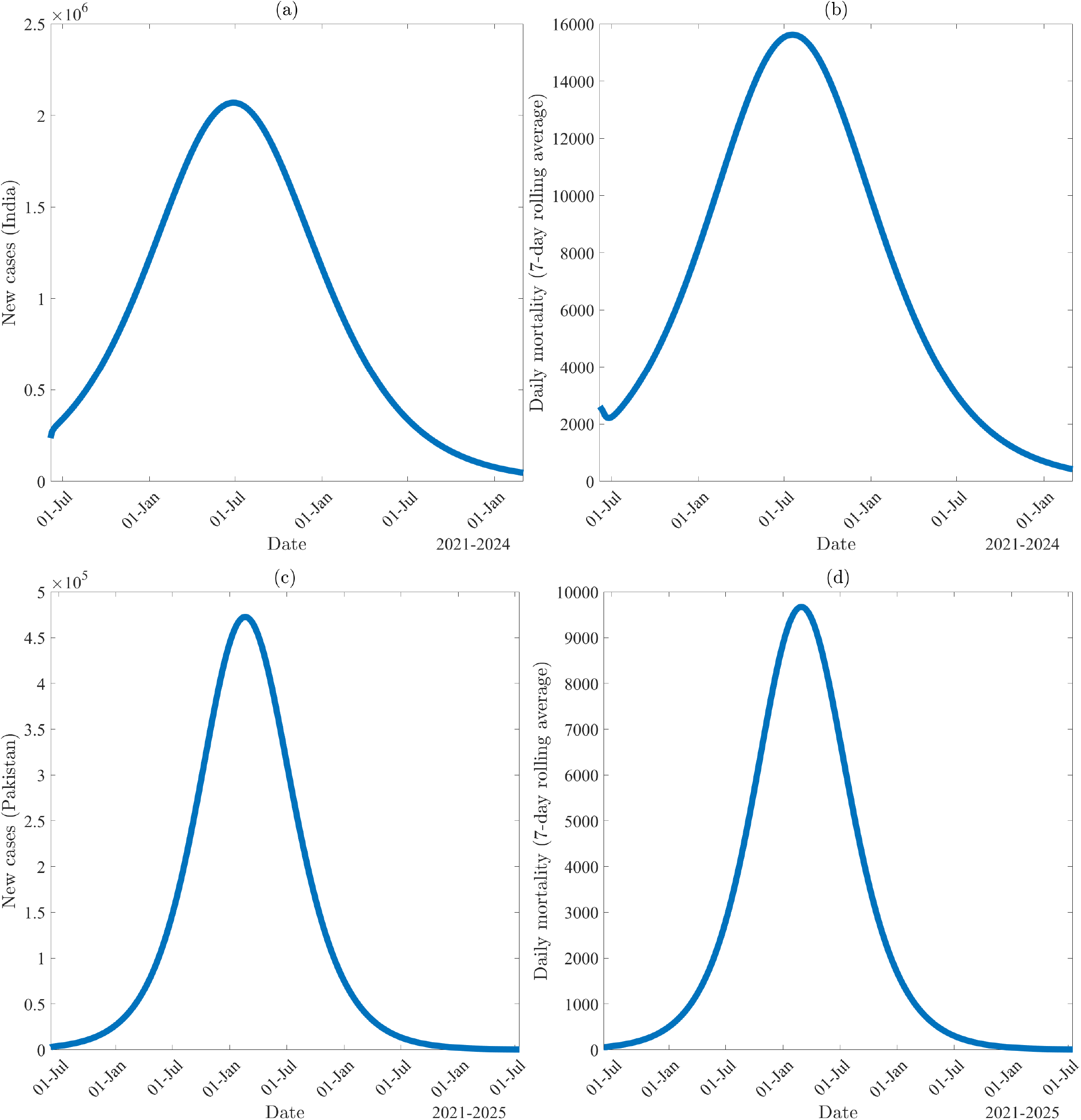
Numerical simulations of the basic model (2.1) for the case where control measures are relaxed to the extent that the baseline levels of the community contact rate parameters (*β*_*P*_, *β*_*I*_, *β*_*A*_, *β*_*H*_) are increased by 40%. (a) New cases in India, (b) the seven-day rolling average of daily mortality in India, (c) new cases in Pakistan, and (d) the seven-day rolling average of daily mortality in Pakistan. Initial conditions were obtained by simulating the basic model using the parameters estimated in the period from March 1, 2021 to June 6, 2021 together with the values of the fixed parameters listed in Table 3. Values of the estimated parameters prior to June 6, 2021 are listed in Table 4a (India) and Table 4b (Pakistan). Values for the estimated parameters after June 6, 2021 are computed by increasing the baseline values of the estimated contact rates (*β*_*P*_, *β*_*I*_, *β*_*A*_, *β*_*H*_) by 40%. Values estimated during the lockdown period are used for other estimated parameters (*ϕ, δ*_*I*_, *δ*_*H*_).

Figure 3(b) depicts the seven-day average daily COVID-19 mortality, as a function of time, for various effectiveness levels of the lockdown measures implemented in India during the second pandemic wave. This figure shows that the second wave of the pandemic in India has peaked around May 20, 2021 (Figure 3(b), red dots). Here, too, strengthening lockdown measures, as measured by a 20% decrease in the baseline values of the community-contact rate parameters, significantly decreases the projected daily mortality (Figure 3(b), purple curve), while relaxation of the lockdown measures (corresponding to a 20% or a 40% increase in the baseline values of the contact rate parameters) resulted in a marked increase in the projected daily mortality (Figure 3(b), brown and blue curves, respectively). In summary, the simulations in Figure 3 suggest that relaxing the current levels of lockdown measures in India could lead to a third wave of the pandemic in the country. The simulation results for the dynamics of the pandemic in India, for the various lockdown relaxation and strengthening scenarios discussed above, are further summarized in Table 5a.

**Table 5:** Estimation of the basic reproduction numbers for India and Pakistan (*R*_0*i*_ and *R*_0*p*_), projected cumulative mortality generated from the basic model (2.1), and changes (both absolute and relative) from the baseline case on September 4, 2021. Parameter values used are as given in Table 3 (for both countries) and Table 4a (for India) and Table 4b (for Pakistan).

| Case | $\mathcal{R}_{0i}$ | Cumulative Deaths (Change) | % Change from Baseline |
| --- | --- | --- | --- |
| No change (baseline) | 0.79 | 459,000 | N/A |
| 20% reduction in $\beta$ | 0.63 | 434,000 (−25,000) | $\approx 5.4\%$ decrease |
| 20% increase in $\beta$ | 0.95 | 507,000 (+48,000) | $\approx 10.5\%$ increase |
| 40% increase in $\beta$ | 1.11 | 599,000 (+140,000) | $\approx 30.5\%$ increase |

| Case | $\mathcal{R}_{0p}$ | Cumulative Deaths (Change) | % Change from Baseline |
| --- | --- | --- | --- |
| No change (baseline) | 0.80 | 24,100 | N/A |
| 20% reduction in $\beta$ | 0.64 | 23,100 (−1,000) | $\approx 4.1\%$ decrease |
| 20% increase in $\beta$ | 0.96 | 26,100 (+2,000) | $\approx 8.3\%$ increase |
| 40% increase in $\beta$ | 1.12 | 30,500 (+6,400) | $\approx 26.6\%$ increase |

Similar simulations are also carried out for the dynamics of the pandemic in Pakistan. Specifically, Figure 3(c) depicts the cumulative COVID-19 mortality for Pakistan, as a function of time, for various levels of relaxation and strengthening of the lockdown measures implemented in Pakistan. This figure shows that Pakistan could record a projected 24, 100 cumulative deaths due to COVID-19 by September 4, 2021, under the baseline scenario (Figure 3(c), gold curve). Under this baseline scenario, the reproduction number for COVID-19 in Pakistan is estimated to be ℛ_0*p*_ *≈* 0.8, suggesting that the COVID-19 epidemic in Pakistan is on the downward trajectory. If the lockdown measures are further strengthened to a level that corresponds to a 20% decrease in the baseline values of the community contact rate parameters, the basic reproduction number for Pakistan further reduces to ℛ_0*p*_ *≈* 0.64 (in addition to averting 1, 000 of the projected cumulative mortality by September 4, 2021), thereby enhancing the prospect of speedily suppressing the pandemic in Pakistan (Figure 3(c), purple curve).

Figure 3(c) further shows that if the lockdown measures are relaxed to a level that corresponds to a 20% increase in the community contact rate parameters, about 2, 000 additional deaths could be recorded by September 4, 2021 (Figure 3(c), brown curve), corresponding to a 8.3% increase in the projected cumulative mortality for Pakistan by September 4, 2021. For this level of increase in community contact rate parameters, the basic reproduction number of the pandemic in Pakistan increases to ℛ_0*p*_ *≈* 0.96, suggesting an upward trajectory to more sustained outbreaks of the pandemic in Pakistan. If the lockdown measures are further relaxed to correspond to a 40% increase in the baseline values of the effective contact rate parameters, our simulations (Figure 3(c), blue curve) show that Pakistan could record an additional 6, 400 deaths by September 4, 2021 (representing a 26.6% increase from the projected baseline). For this relaxation scenario, the basic reproduction number increases to ℛ_0*p*_ *≈* 1.12, suggesting that Pakistan could experience a fourth wave of the pandemic (which would be relatively mild). Under this lockdown relaxing scenario, the predicted fourth wave of the pandemic in Pakistan could peak by mid-February 2023 (Figure 4(c)), while the daily mortality could peak near the end of the same month (Figure 4(d)).

Time-to-elimination projections for India and Pakistan, under various levels of control measures, are tabulated in Table 6. Time-to-elimination is defined as the date when new cases of COVID-19 are asymptotically zero. Under the current lockdown implementation, COVID-19 is projected to be eliminated in July 2022 in India and February 2022 in Pakistan. If lockdown measures are strengthened such that community contact rates decrease by 20%, these projections are shortened to December 2021 in India, and September 2021 in Pakistan. However, if control measures are relaxed such that the community contact rate parameters are increased by 20% (from their baseline levels), our simulations show that the pandemic can be eliminated in India and Pakistan by September and October of 2025, respectively. Elimination time is further extended if lockdown measures are relaxed to a level corresponding to a 40% increase in baseline values of the community contact rate parameters. Overall, our simulations show that relaxing the current levels of the control and mitigation measures implemented in the two countries (as measured in terms of increases in the corresponding community contact rate parameters from their current baseline values) prolongs the time-to-elimination of the pandemic.

**Table 6:** Estimated time-to-elimination of COVID-19 in India and Pakistan using the basic model (2.1) under various effectiveness levels of lockdown. Time-to-elimination is measured as the time when new infections are zero asymptotically.

|  | India |  | Pakistan |  |
| --- | --- | --- | --- | --- |
| Case | $\mathcal{R}_{0i}$ | Date | $\mathcal{R}_{0p}$ | Date |
| No change (baseline) | 0.79 | July 2022 | 0.80 | Feb. 2022 |
| 20% reduction in $\beta$ | 0.63 | Dec. 2021 | 0.64 | Sep. 2021 |
| 20% increase in $\beta$ | 0.95 | Sep. 2025 | 0.96 | Oct. 2025 |
| 40% increase in $\beta$ | 1.11 | Nov. 2026 | 1.12 | June 2026 |

Simulations for the seven-day averages of the COVID-19 mortality in Pakistan, as a function of time, under the aforementioned relaxation and strengthening levels of the lockdown measures implemented in Pakistan, are depicted in Figure 3(d). This figure shows that the daily mortality appears to have peaked around May 1, 2021 (red dots in Figure 3(d)). If the lockdown measures are strengthened to correspond to a 20% reduction in the baseline values of the community contact rate parameters, our simulations show a mild reduction in the projected cumulative mortality (Figure 3(d), purple curve). However, if the lockdown measures are relaxed to the extent that the baseline community contact rate parameters are increased by 20%, our simulations suggest a generally mild increase in the projected cumulative mortality by September 4, 2021 (Figure 3(d), brown curve). Furthermore, if such relaxation resulted in a 40% increase in the baseline levels of the community contact rate parameters, a more severe epidemic will result (Figure 3(d), blue curve). The simulation results for the dynamics of the pandemic in Pakistan are summarized in Table 5b. Similar to our projections for India, these simulations show that relaxing the lockdown measures, at the aforementioned moderate to high levels (in comparison to the current baseline levels), may trigger a fourth wave of the COVID-19 pandemic in Pakistan.

Having analysed the dynamics of the pandemic in each of the two countries individually, it is instructive to assess the impact of travels between the two countries on the trajectory of the pandemic in each of the two countries. To achieve this objective, the basic model (2.1) will now be extended to include the back-and-forth mobility between the two nations.

## 3 Metapopulation Model for COVID-19 Dynamics in India and Pakistan

India (population over 1.38 billion) and Pakistan (population over 220 million) are geographical neighbours, sharing a 2,000 kilometer (1,243 mile) border, with an additional 1,300 kilometers (808 miles) disputed [55, 56]. The volume of travel between India and Pakistan, during non-pandemic time, is generally moderate. Since the suspension of the “friendship bus” (the symbolic public transportation link between the two countries), travel between the two nations is limited to air transportation and/or crossing the only remaining open border at Wagah by foot [57]. During the year 2018, India received over 41,000 visitors from Pakistan, down from 100,000 in 2016 [58]. In 2016, over 48,000 residents of India travelled to Pakistan [59]. Although additional travel restrictions between the two countries have been imposed since the emergence of the COVID-19 pandemic [60], it is instructive to study the impact of the back-and-forth mobility between the two countries on the transmission dynamics and control of the pandemic in the two countries. This is particularly relevant considering the fact that India has become the global epicenter of COVID-19 in the spring of 2021.

To achieve the above objective, the basic model (2.1) will be extended to account for the back-and-forth mobility between the two countries. We assume that the mobility pattern between the two countries is largely *Lagrangian* in nature (i.e., it is for a limited period of time, and not permanent migration) [45]. In order to extend the basic model (2.1) to account for the Lagrangian mobility, we consider each of the two countries to be a single patch. Let the subscripts *i* and *p* represent the residents in the patch for India and Pakistan, respectively. Furthermore, for each patch *k*, we stratify the total population at time *t*, denoted by *N*_*k*_(*t*), into the mutually-exclusive compartments of susceptible (*S*_*k*_), exposed/latent (*E*_*k*_), pre-symptomatic (*P*_*k*_), symptomatic (*I*_*k*_), asymptomatic (*A*_*k*_), hospitalized (*H*_*k*_), and recovered (*R*_*k*_) individuals, so that:

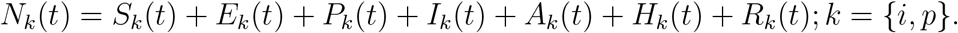

The lowercase indexing notation, *k* = {*i, p}*, is not to be confused with the uppercase indexing notation *P* and *I*, which refer to the populations of individuals in the pre-symptomatic (*P*) and symptomatic (*I*) infectious compartments, respectively.

Let *β*_*P,k*_, *β*_*I,k*_, *β*_*A,k*_, and *β*_*H,k*_ represent, respectively, the effective contact rates for pre-symptomatic, symptomatic, asymptomatic, and hospitalized infectious individuals in patch *k*. The next step in the formulation of the extended model is to track the amount of time individuals spend in patch *k* (*residence-time*).

### 3.1 Formulation of Functional Forms of Residence-Time

Let 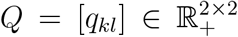 represent the residence-time matrix where *q*_*kl*_ = {*q*_*ii*_, *q*_*ip*_, *q*_*pi*_, *q*_*pp*_*}*. Furthermore, we assume that an individual’s decision to visit a particular patch (during the time of the pandemic) is dependent on the daily number of deaths due to the pandemic in the resident and destination patches. Let 𝒟_*i*_(*t*) and 𝒟_*p*_(*t*) represent the daily COVID-19 deaths in India and Pakistan at time *t*, respectively. Hence, we define

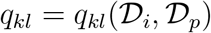

to be the mortality-dependent proportion of time a resident of patch *k* spends in patch *l*. Although Lee *et al*. [45] defined the residence-time matrix *q*_*kl*_ as a function of the number of new COVID-19 infections reported daily in each of the two patches, daily mortality is chosen instead due to the likely under-counting of cases (although deaths may be under-reported as well) [61, 62]. We assume individuals are less likely to choose to travel to a patch with high daily COVID-19 mortality, and that those living in a patch with higher daily COVID-19 mortality may opt to go to a patch with lower daily mortality. To account for these assumptions, the following inequalities must hold [46]:

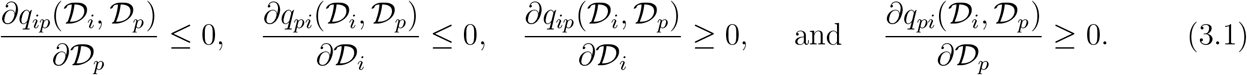

Differentiating *q*_*kk*_ + *q*_*kl*_ = 1 (for *k* ≠*l*), with respect to *𝒟*_*i*_ and *𝒟*_*p*_, reduces the inequalities in (3.1) to:

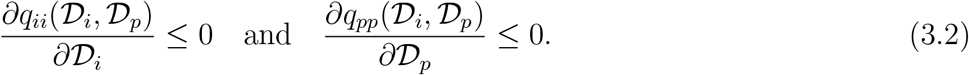

Based on the above assumptions and analyses, we choose the following functional forms for *q*_*kl*_ (*k, l* = {*i, p}*), which satisfy the two inequalities in (3.2):

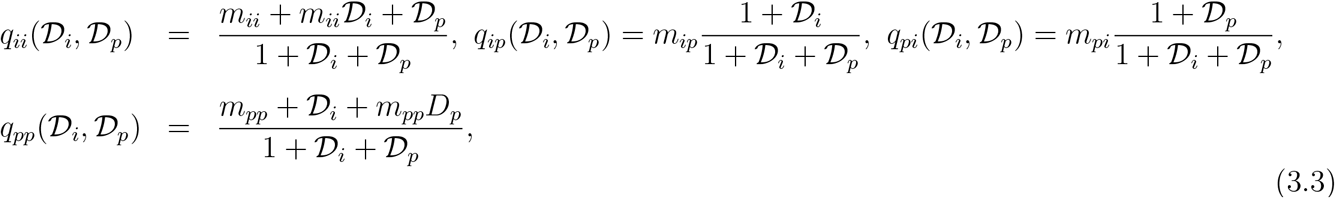

with *m*_*ii*_ + *m*_*ip*_ = 1 and *m*_*pp*_ + *m*_*pi*_ = 1. It can be seen from (3.3) that, if the daily COVID-19 mortality of the two patches is negligible, such that *𝒟*_*i*_ = *𝒟*_*p*_ *≈* 0), the quantity *q*_*kl*_ reduces to the constants *m*_*kl*_ (where *k, l* = *{i, p}*). That is, if the disease does not cause significant mortality in any of the two patches, then *q*_*kl*_ is constant (maintained at the baseline level of proportion of time spent in the patches). Table 8 depicts the values of *m*_*kl*_ used in numerical simulations.

### 3.2 Keeping Track of Total Residents in Each Patch

To keep track of the total number of residents in each of the two patches (countries) at time *t*, we let *M*_*kl*_ represent the total number of residents of patch *k* who are now in patch *l* (see Table 7 for the four possibilities for *M*_*kl*_).

**Table 7:** Total number of residents of patch *k* in patch *l* at time *t* (*M*_*kl*_; *k, l* = *{i, p}*). Subscripts *i* and *p* represent the patch for India and Pakistan, respectively.

| Total Variable | Description |
| --- | --- |
| $M_{ip}(t)$ | Total number of residents of India in Pakistan at time $t$ |
| $M_{ii}(t)$ | Total number of residents of India in India at time $t$ |
| $M_{pi}(t)$ | Total number of residents of Pakistan in India at time $t$ |
| $M_{pp}(t)$ | Total number of residents of Pakistan in Pakistan at time $t$ |

We assume that everyone but symptomatic (*I*_*k*_) and hospitalized (*H*_*k*_) residents of patch *k* can travel to the other patch. Hence, it follows from Table 7 that (recalling that *N*_*i*_(*t*) is the total number of residents of India at time *t*):

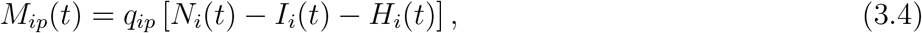

where *N*_*i*_(*t*) *− I*_*i*_(*t*) *− H*_*i*_(*t*) is the total number of residents of India who can travel (to Pakistan) at any time *t*. Furthermore,

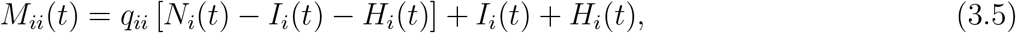

where *q*_*ii*_ [*N*_*i*_(*t*) *− I*_*i*_(*t*) *− H*_*i*_(*t*)] is the total number of residents of India who can travel (but choose to remain in India), while *I*_*i*_ + *H*_*i*_ is the total number of symptomatic and hospitalized residents of India who cannot travel outside of India. Similarly, *M*_*pi*_ and *M*_*pp*_ are given, respectively, by:

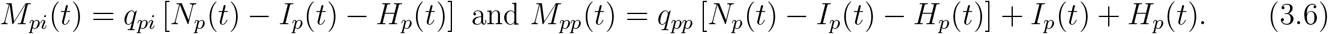

It should be noted from Equations (3.4)-(3.6) that:

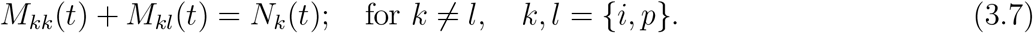

The conservation law described in (3.7) implies that the total number of residents of patch *k* (with *k* = *{i, p}*) at any time *t* is the sum of the total number of residents of patch *k* residing in patch *k* at time *t* (*M*_*kk*_(*t*)) and the total number of residents of patch *k* currently in patch *l* at time *t* (*M*_*kl*_(*t*)). That is, the sum *M*_*kk*_(*t*) + *M*_*kl*_(*t*) is total number of residents of a patch *k* at time *t*, while the sum *M*_*kk*_(*t*) + *M*_*lk*_(*t*) represents the total number of people located in patch *k* at time *t*.

### 3.3 Keeping Track of Infection Rates

Susceptible individuals from patch *k* = {*i, p}* can be infected (by infectious individuals) *via* four main transmission pathways. For example, susceptible individuals from India can acquire COVID-19 infection while residing in India or while visiting Pakistan. Specifically, susceptible residents of India currently in India (*q*_*ii*_*S*_*i*_(*t*)) can acquire COVID-19 infection from infectious individuals who are (i) residents of India who are currently in India or (ii) who are residents of Pakistan but currently visiting India. Similarly, susceptible residents of India who are currently in Pakistan (*q*_*ip*_*S*_*i*_(*t*)) can acquire COVID-19 infection from infectious individuals who are (i) residents of India but currently visiting Pakistan or (ii) residents of Pakistan who are currently in Pakistan.

Let *λ*_*kk*_ be the rate at which residents of patch *k* acquire infection in patch *k*. Hence,

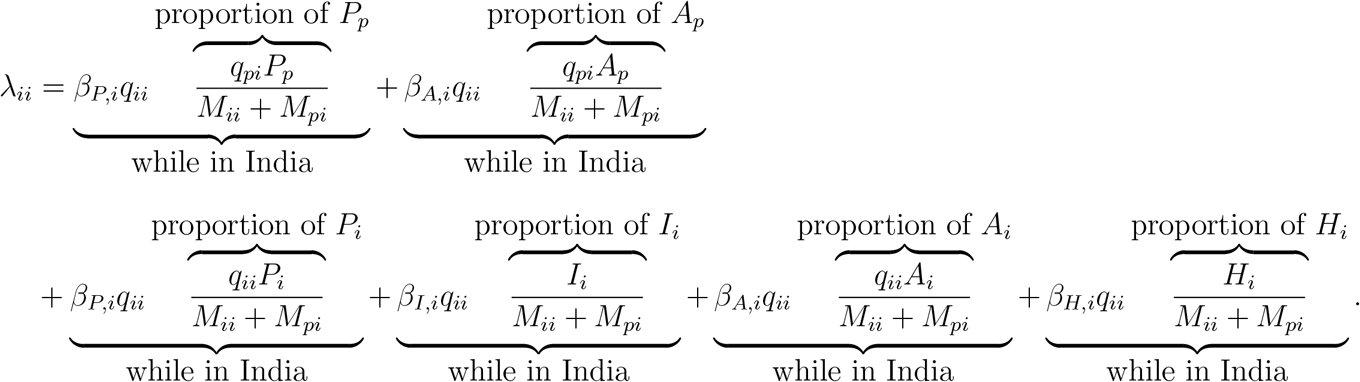

Similarly,

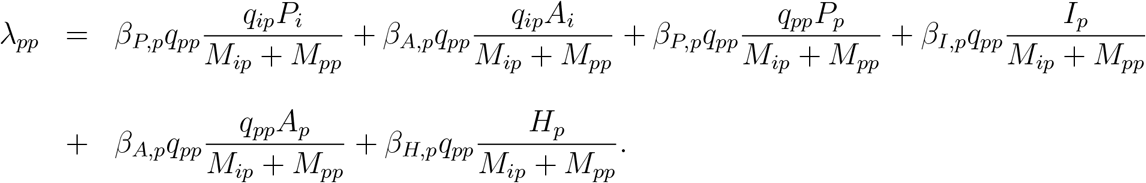

Furthermore, let *λ*_*kl*_ represent the rate at which residents of patch *k* acquire infection while in a different patch *l*. Thus,

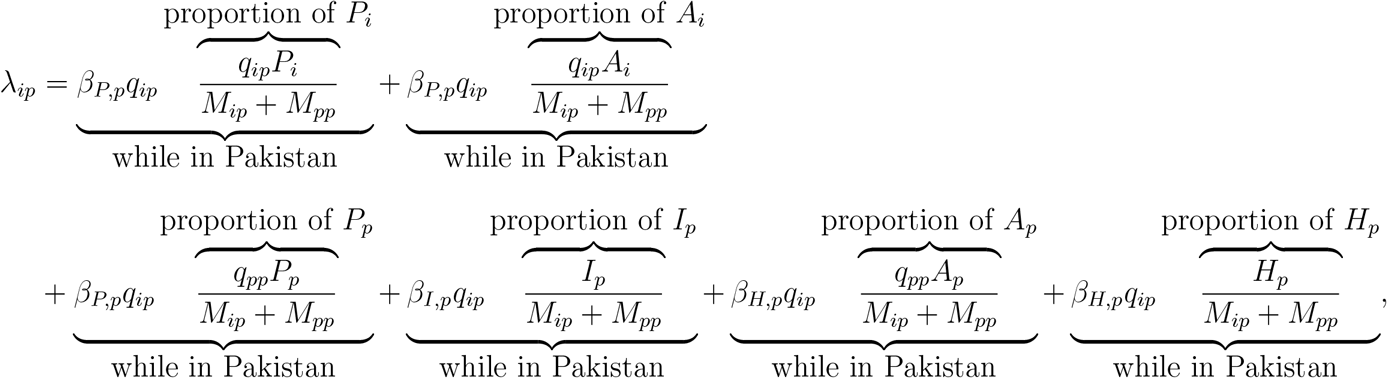

and,

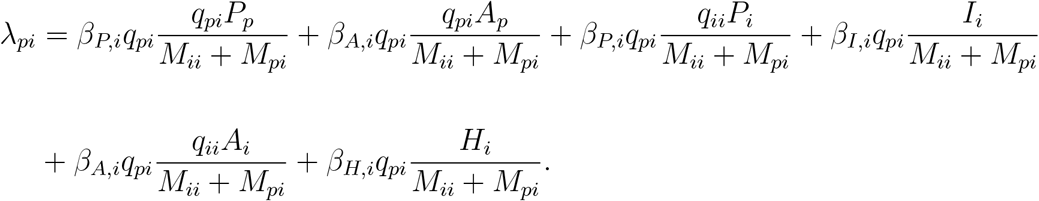

### 3.4 Equations of Metapopulation Model

It follows from the above formulation, derivation, and assumptions that the metapopulation model for the transmission dynamics of COVID-19 between India and Pakistan, which incorporates Lagrangian mobility, is given by the following deterministic system of nonlinear-differential equations (a flow diagram of the metapopulation model (3.8) is depicted in Figure 5 and the state variables and parameters of the model are described in Tables 1 and 2, respectively).

**Figure 5:**
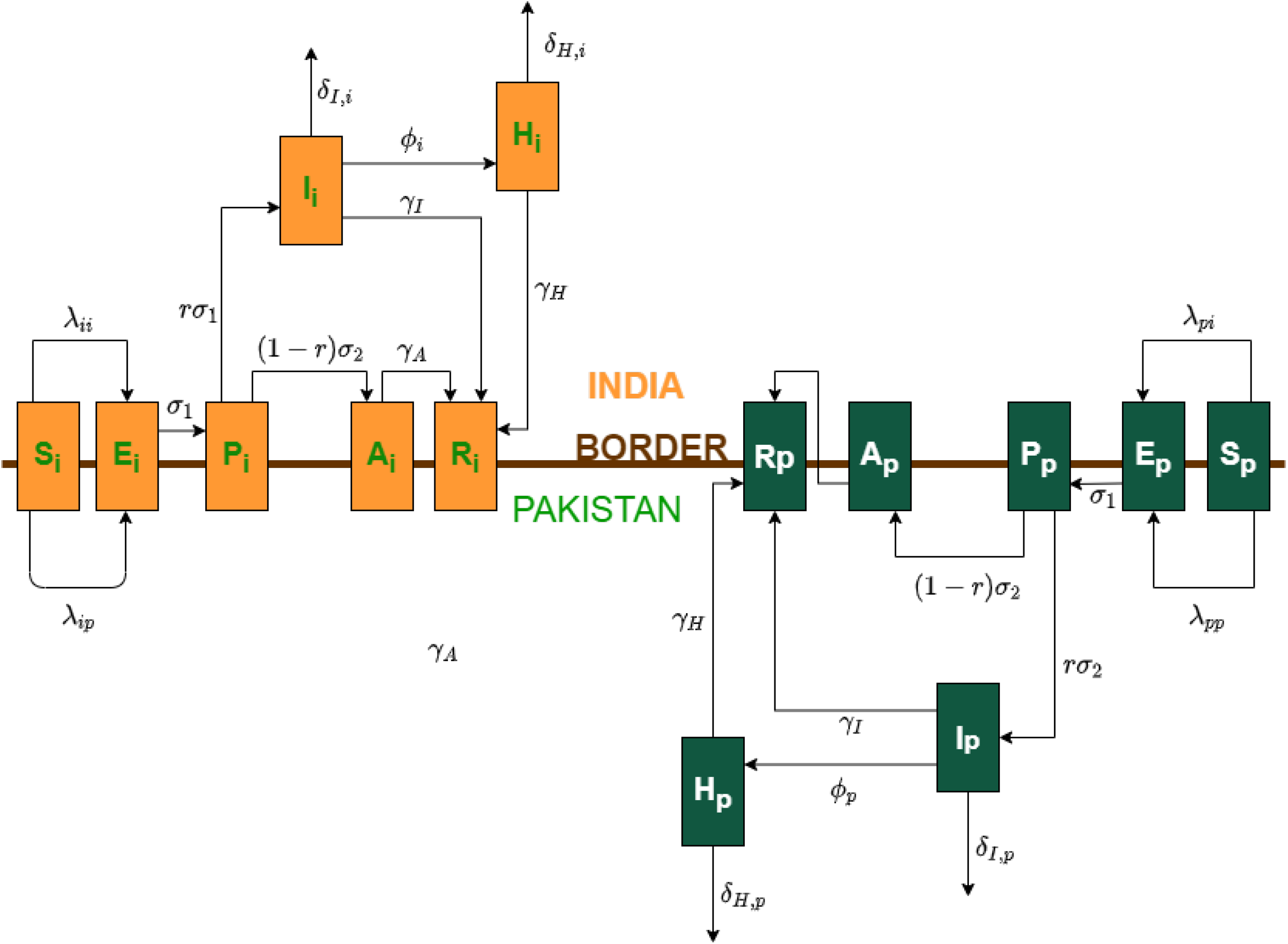
Flow diagram of the metapopulation model (3.8). Gold and green boxes represent compartments of the metapopulation model corresponding to the India and Pakistan patch, respectively. Populations of susceptible (*S*), exposed (*E*), pre-symptomatic (*P*), asymptomatic (*A*), and recovered (*R*) compartments can travel between the two countries (hence, they are placed along the hypothetical “border” of the two nations). Symptomatically-infectious (*I*) and hospitalized individuals (*H*) cannot travel due to their ill health (hence, they remain in their respective patches).

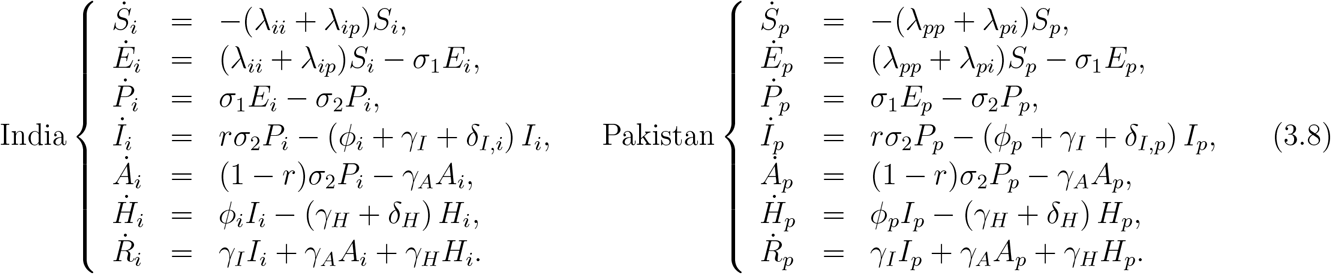

For bookkeeping purposes, we let *D*_*k*_(*t*) represent the number of COVID-19 mortality in of residents of patch *k* at time *t* (with *k* = *i, p*). Hence, the rate of change of the population *D*_*k*_ is given by:

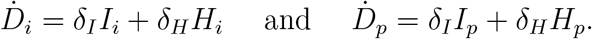

The metapopulation model (3.8) is an extension of the COVID-19 patch model in [45] by, *inter alia*:

i. adding an exposed/latent compartment (*E*_*k*_);
ii. splitting the transmission-capable individuals in patch *k* into four compartments, namely the pre-symptomatic (*P*_*k*_), symptomatic (*I*_*k*_), asymptomatic (*A*_*k*_), and hospitalized (*H*_*k*_) compartments (as opposed to a single infectious class);
iii. redefining the residence-time matrix *q*_*kl*_ with *k, l* = *{i, p}* to be dependent on daily deaths in each patch (𝒟_*i*_ and 𝒟_*p*_), as opposed to number of daily infections due to the likely under-counting of COVID-19 cases [61];
iv. excluding the possibility of sick and hospitalized individuals from travelling between patches. In particular, we assume that symptomatic (*I*_*k*_) and hospitalized (*H*_*k*_) residents of patch *k* do not (or cannot) travel to the other patch *l*, in contrast to all infectious individuals being able to travel between patches.

#### 3.4.1 Basic reproduction number for metapopulation model

The derivation of the (overall) reproduction number of the metapopulation model (denoted by *ℛ*_0*m*_), using the next generation operator method, is presented in Appendix C. These analyses reveal that *ℛ*_0*m*_ is expressed as the maximum of the constituent reproduction numbers for the residents of India (*ℛ*_0*ri*_) and Pakistan (*ℛ*_0*rp*_). That is,

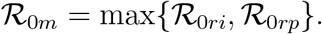

Figure 6 depicts contour plots of the reproduction number of the metapopulation model (3.8), as a function of the proportion of the residence-time residents of one country spend in the other country, under various lockdown scenarios. Specifically, Figure 6(a) depicts a contour of the reproduction number generated using the baseline lockdown scenario (where the baseline values of the estimated parameters listed under the “during lockdown” columns in Table 4, as well as the fixed parameters in Table 3, are used). This figure shows that the overall reproduction of the metapopulation model lies between 0.78 to 0.83, and that ℛ_0*m*_ mildly increases with increasing time residents of India spend in Pakistan (and decreases with increasing time residents of Pakistan spend in India). This result is intuitive considering the fact that India is experiencing a much larger epidemic compared to Pakistan, and increased mobility from India to Pakistan will cause more cases in Pakistan (i.e., the virus presumably has more hosts to infect in Pakistan, compared to in India, where the level of natural immunity may be significant).

**Figure 6:**
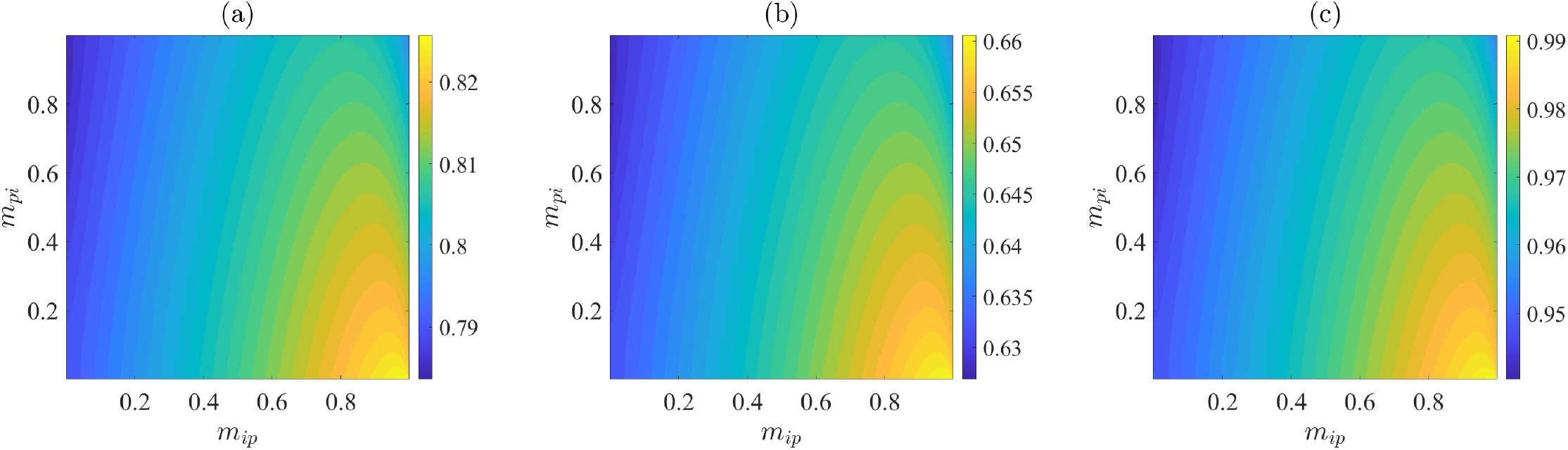
Contour plots of the overall reproduction number (*ℛ*_0*m*_) of the metapopulation model (3.8), as a function of resident-times residents of one country spend in the other country (*m*_*ip*_ and *m*_*pi*_). (a) Baseline lockdown scenario (parameter values used as listed under the “during lockdown” columns in Table 4, together with the fixed parameters tabulated in Table 3). (b) Lockdown measures are further strengthened to the extent that the baseline values of the community contact rate parameters (*β*_*P*_, *β*_*I*_, *β*_*A*_, *β*_*H*_) are decreased by 20%. (c) Lockdown measures are mildly relaxed to the extent that the community contact rate parameters are increased by 20%. The reproduction number is computed using the next generation method in Appendix C. Values for fixed parameters for both countries are listed in Table 3. Values for parameters estimated during lockdown (used as the baseline) are listed in Table 4a (for India) and Table 4b (for Pakistan).

If lockdown measures are slightly increased to the extent that the baseline values of the contact rate parameters of the metapopulation model (*β*_*P*_, *β*_*I*_, *β*_*A*_, *β*_*H*_) are reduced by 20%, our simulations (Figure 6 (b)) show a decrease in the range of ℛ_0*m*_ to now fall between 0.62 to 0.67. However, if lockdown measures are relaxed to the extent that the baseline values of the community contact rate parameters are increased by 20%, the range of ℛ_0*m*_ increases to lie between 0.94 to 0.99 (Figure 6 (c)). In summary, the contour plots depicted in Figure 6 show a more promising prospect of the pandemic in both countries under the current baseline scenario, since the values of the overall reproduction number (ℛ_0*m*_) generally lie consistently below one (even for the case when lockdown measures are mildly relaxed to generate a 20% increase in the baseline levels of the community transmission parameters, although *ℛ*_0*m*_ is near unity). These plots further show that increase in mobility from India to Pakistan causes a corresponding increase in ℛ_0*m*_ (i.e., causes an overall increase in disease burden in both countries), and increase in mobility in the opposite direction causes a mild decrease in overall disease burden.

It is worth mentioning that, for the case where no mobility between the patches is allowed (i.e., *m*_*pi*_ = *m*_*ip*_ = 0), the metapopulation model (3.8) reduces to the single-patch model (2.1). Under the baseline scenario described above, the reproduction number of the single-patch model for India (*ℛ*_0*i*_) and Pakistan (*ℛ*_0*p*_) is 0.79 and 0.8, respectively (and the reproduction number of the metapopulation model (*ℛ*_0*m*_) is the maximum of the reproduction number for Pakistan and India, namely *ℛ*_0*m*_ = *ℛ*_0*p*_ = 0.8).

#### 3.4.2 Numerical simulations

The metapopulation model (3.8) will now be simulated to assess the impact of the back-and-forth mobility between India and Pakistan on the transmission dynamics of the COVID-19 pandemic in each of the two nations. Specifically, we will run the simulations from the period June 6, 2021 (when lockdown measures are generally partially- or fully-lifted in most regions within the two countries) and make predictions for a 90-day period (i.e., until September 4, 2021). That is, we consider the total residence-time to be 90 days. It is worth stating that, despite the stringent lockdown and travel restriction measures that may have been implemented in jurisdictions within the two nations (particularly during the height of the last devastating wave of the pandemic), a certain level of of travel between the nations still occurs, largely for economic reasons [34].

For the simulations to be carried out, the initial conditions (of the state variables) for the metapopulation model (3.8) were obtained from simulating the basic model (2.1) (using the fixed and estimated parameter values, both for before and during lockdowns, given in Section 2.3) for the period March 1, 2021 to June 6, 2021. Similarly, the values of the the residence-time proportions, *m*_*kl*_ (which represents the proportion of time a resident of patch (or country) *k* spends in patch (or country) *l*), used in the simulations of the metapopulation model are tabulated in Table 8. For simulation purposes, we consider the following four mobility/dispersal scenarios (tabulated in Table 8), depending on the preference to stay in one patch, in relation to the other):

a. Semi-symmetric 1: here, individuals of each patch spend 99% of their residence-time (i.e., 89.1 days) in their own patch and only 1% (i.e., approximately 0.9 days) in the other. Consequently, we set *m*_*ii*_ = *m*_*pp*_ = 0.99 and *m*_*ip*_ = *m*_*pi*_ = 0.01. Since simulations are carried out for 90 days, *m*_*ip*_ = *m*_*pi*_ = 0.01 implies that residents spend approximately *m*_*ip*_ *×* 90 or 0.9 days in the other patch.
b. Semi-symmetric 2: under this scenario, individuals spend 95% of their residence-time (i.e., 85.5 days) in their own patch and 5% (or 4.5 days) in the other, so that *m*_*ii*_ = *m*_*pp*_ = 0.95 and *m*_*ip*_ = *m*_*pi*_ = 0.05.
c. Uni-directional 1: here, we consider the scenario where residents of Pakistan do not travel to India and residents of India spend 1% of their residence-time in Pakistan. We set *m*_*ii*_ = 0.99, *m*_*pp*_ = 1, *m*_*ip*_ = 0.01 and *m*_*pi*_ = 0. This is a reasonable scenario to explore, particularly during the height of the third wave of the pandemic when India became the global epicenter of the COVID-19 pandemic (hence, residents of Pakistan, who was experiencing a much milder outbreak, would opt to stay in their own patch, rather than travelling to India). Furthermore, residents of a nation that is experiencing a major outbreak may consider it reasonable to travel to a patch experiencing milder or no outbreak at all.
d. Uni-directional 2: for this scenario, residents of India do not travel to Pakistan but residents of Pakistan can spend 1% of their residence-time in India. Thus, *m*_*ii*_ = 1, *m*_*ip*_ = 0, *m*_*pp*_ = 0.99 and *m*_*pi*_ = 0.01. This scenario is particularly motivated by the fact that, on April 19, 2021, Pakistan banned almost all non-Pakistani travellers from India to enter Pakistan by land or air [63, 64].

**Table 8:** Proportions of residence-time (*m*_*kl*_) for the metapopulation model (3.8), where *m*_*kl*_ denotes the proportion of time residents of patch *k* spend in patch *l* (the values of *m*_*kl*_ given in this table are used in the behavioral response equations given in (3.3)). The subscripts *i* and *p* denote the patch for India and Pakistan, respectively.

| Mobility Scenario | Proportion of Residence-Time |
| --- | --- |
| Semi-symmetric 1 | $m_{ii} = m_{pp} = 0.99; m_{ip} = m_{pi} = 0.01$ |
| Semi-symmetric 2 | $m_{ii} = m_{pp} = 0.95; m_{ip} = m_{pi} = 0.05$ |
| Uni-directional 1 | $m_{ii} = 0.99, m_{pp} = 1; m_{ip} = 0.01, m_{pi} = 0$ |
| Uni-directional 2 | $m_{ii} = 1, m_{pp} = 0.99; m_{ip} = 0, m_{pi} = 0.01$ |

Further, we explore the mobility scenarios under three levels of lockdown effectiveness, as measured in terms of increases or decreases in the baseline values of the community contact rate parameters (*β*_*P*_, *β*_*I*_, *β*_*A*_, *β*_*H*_), namely:

i. Maintaining baseline level of lockdown measures: in this case, baseline values of both the fixed and estimated parameters of the metapopulation model, for the lockdown period (given in Section 2.3, for the period March 1, 2021 to June 6, 2021), will be used.
ii. Strengthening of lockdown measures: here, the lockdown measures implemented in the two patches are enhanced (by achieving increased compliance and effective implementation of other contact-reduction measures) such that the baseline levels of the community contact rate parameters (during lockdown) are decreased by 20%.
iii. Relaxation of lockdown measures: in this scenario, the lockdown measures are relaxed to the extent that the community contact rate parameters are increased by a factor of 20% from their baseline values during lockdown.

Figure 7 depicts the cumulative COVID-19 mortality for India (a-b, e-f) and Pakistan (c-d, g-h), as a function of time, under the two semi-symmetric mobility scenarios, for various levels of lockdown. This figure shows that, under the baseline and semi-symmetric 1 scenario (Figures 7(a) and (c), blue curves), India and Pakistan are projected to record approximately 459, 000 and 26, 300 cumulative deaths by September 4, 2021, respectively (the corresponding reproduction number for the metapopulation model is ℛ_0*m*_ *≈* 0.79, suggesting that the pandemic will be effectively suppressed in both patches if the baseline levels of the lockdown measures are maintained and if mobility between the two nations follows this semi-symmetric modality). The seven-day rolling-average for daily mortality under this mobility scenario for India and Pakistan is depicted in Figures 7(b) and (d), respectively. Under baseline lockdown levels, daily mortality in India decreases over time (Figure 7(b), blue curve). On the other hand, the daily mortality in Pakistan increases slightly, peaks near the end of June 2021, and then decreases (Figure 7(d), blue curve).

**Figure 7:**
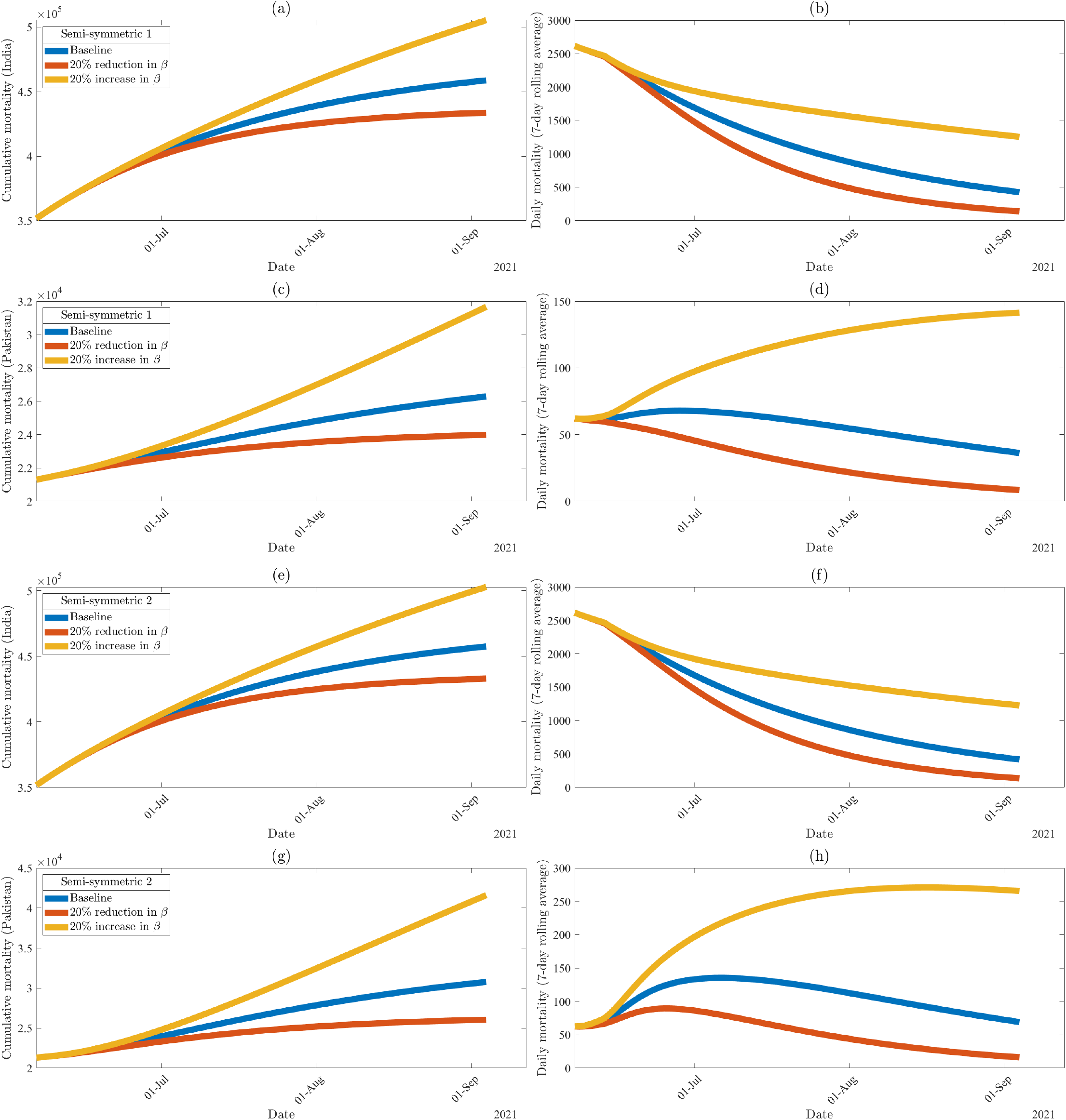
Numerical simulations of the two-patch metapopulation model (3.8) for various effectiveness levels of the implementation or relaxation of community lockdown measures under the semi-symmetric mobility scenarios listed in Table 8, showing (a-b) cumulative and seven-day rolling average daily mortality projections for India and under semi-symmetric 1, (c-d) cumulative and seven-day rolling average daily mortality projections for Pakistan and under semi-symmetric 1, (e-f) cumulative and seven-day rolling average daily mortality projections for India and under semi-symmetric 2, and (g-h) cumulative and seven-day rolling average daily mortality projections for Pakistan and under semi-symmetric 2. Initial conditions were obtained via simulation of the basic model (2.1) up to June 6, 2021. Values for fixed parameters for both countries are listed in Table 3. Values for parameters specific to each country during lockdown are listed in Table 4a (India) and Table 4b (Pakistan). Effectiveness of lockdown is measured by increasing or decreasing the estimated community contact rates (*β*_*P*_, *β*_*I*_, *β*_*A*_, *β*_*H*_) by the indicated percentage. Values obtained during the lockdown period for each country are used for other estimated parameters (*ϕ, δ*_*I*_, *δ*_*H*_).

Furthermore, for this semi-symmetric 1 scenario, if the lockdown measures are further strengthened from their baseline levels, such that the community contact rate parameters are decreased by 20%, our simulation results (Figures 7(a) and (c), brown curves) show a decrease in the projected cumulative mortality for India and Pakistan to be 434, 000 and 24, 000, respectively. Thus, strengthening lockdown measures in India and Pakistan (from June 6, 2021 to September 4, 2021) to the level that resulted in a 20% decrease in baseline values of the community contact rate parameters would have averted approximately 25, 000 and 2, 300 projected cumulative deaths by September 4, 2021 (this corresponds to 5.4% and 8.7% reduction in the projected cumulative mortality for the two nations, respectively; for this scenario, the reproduction number decreases to 0.63, further suggesting an improved likelihood of pandemic elimination in both countries). Here, daily mortality in both countries declines steadily (Figures 7(b) and (d), brown curves).

On the other hand, if the lockdown measures are relaxed, corresponding to a 20% increase in the baseline values of the community contact rate parameters, our results depicted in (Figures 7(a) and (c), gold curves) show that while India will record a projected additional 46, 000 deaths (a 10% increase from the projected value under the baseline scenario), Pakistan will also record an additional 5, 400 deaths (representing a 20.5% increase in comparison to the projected number under the baseline scenario). For this scenario, where lockdown measures are relaxed to correspond to a 20% increase in the contact rate parameters, the reproduction number of the metapopulation model increases to 0.95. Thus, this result shows that the pandemic can still be mitigated in the two countries even if the lockdown measures implemented in the countries are mildly lifted (to correspond to a 20% increase in the baseline values of the contact rate parameters), but the major downside to this relaxation is that it will result in significant number of deaths than can be averted if such relaxations were not implemented. While the daily mortality in India declines more slowly in comparison to more stringent lockdown scenarios (Figure 7(b), gold curve), the daily mortality in Pakistan continues to increase past September 4, 2021 (Figure 7(d), gold curve), indicating the potential for a fourth wave of the pandemic in Pakistan (that would peak in mid-September of 2021).

Cumulative mortality projections under the second semi-symmetric mobility scenario (as a function of time) are shown in Figures 7(e) and (g). Recall that, in semi-symmetric 2, residents of a patch spend approximately 4.5 days in the other patch. Although mortality projections for India do not change significantly when compared to semi-symmetric 1 (458, 000 versus 459, 000 deaths, respectively), the projected cumulative mortality for Pakistan increases to 30, 800 deaths (representing a 17.1% increase from the baseline, in comparison to the result obtained for the semi-symmetric 1 scenario). The associated reproduction number for the baseline semi-symmetric 2 case is *ℛ*_0*m*_ *≈* 0.79. As stated earlier, these simulations show an increase in overall disease burden with increasing values of *m*_*ip*_ (and the increase in burden, measured in terms of disease-induced mortality, is more pronounced in Pakistan than in India).

If lockdown measures are strengthened, corresponding to a 20% decrease in community contact rate parameters, the simulation results obtained (Figures 7(e) and (g), brown curves) show that up to 25, 000 and 4, 800 deaths could be averted in India and Pakistan, respectively, by September 4, 2021 (and *ℛ*_0*m*_ reduces to approximately 0.63). If lockdown measures are relaxed (as discussed above), ℛ_0*m*_ increases to 0.95 and additional 45, 000 and 10, 800 cumulative deaths are projected for India and Pakistan, respectively (Figures 7(e) and (g), gold curves). In this mobility scenario, daily mortality in India declines regardless of lockdown level (Figure 7(f)). In Pakistan, daily mortality increases slightly before decreasing again, with a lower peak occurring earlier when control measures are strengthened (Figure 7(h), brown curve). Similar to the first semi-symmetric scenario, a fourth wave of the pandemic in Pakistan occurs in this scenario when lockdown measures are relaxed (Figure 7(h), gold curve), with peak daily deaths projected to occur in mid-August of 2021.

We now consider the uni-directional mobility scenarios, where residents of one patch stay in their home patch (and, thus, do not travel), while residents of the other patch are able to move between the two patches. Specifically, residents who are able to travel spend 1% of their residence time proportion in the other patch. Figure 8 shows cumulative COVID-19 mortality, as a function of time, under the two unidirectional dispersal scenarios. Our results for the uni-directional 1 mobility scenario show that, under the baseline lockdown level, India will record a projected 459, 000 deaths by September 4, 2021, while Pakistan will record a projected 26, 300 deaths by the same time period (Figure 8(a), blue curves). The overall results obtained for these scenarios are qualitatively very similar to those obtained for the semi-symmetric 1 case (see Table 9). Specifically, we show that relaxing the control and mitigation measures, coupled with increased mobility from India to Pakistan, would trigger a fourth wave of the pandemic in Pakistan. Under these conditions, daily COVID-19-induced mortality would peak in Pakistan around mid-September of 2021.

**Table 9:** Projected cumulative mortality for India and Pakistan generated using the metapopulation model (3.8) under various mobility scenarios and effectiveness levels of lockdown measures. The mobility scenarios are described in Table 8.

| Mobility Scenario | Baseline | 20% reduction in $\beta$ | 20% increase in $\beta$ |
| --- | --- | --- | --- |
| Semi-symmetric 1 | 459,000 | 434,000 | 505,000 |
| Semi-symmetric 2 | 458,000 | 433,000 | 503,000 |
| Uni-directional 1 | 459,000 | 434,000 | 505,000 |
| Uni-directional 2 | 459,000 | 434,000 | 507,000 |

| Mobility Scenario | Baseline | 20% reduction in $\beta$ | 20% increase in $\beta$ |
| --- | --- | --- | --- |
| Semi-symmetric 1 | 26,300 | 24,000 | 31,700 |
| Semi-symmetric 2 | 30,800 | 26,000 | 41,600 |
| Uni-directional 1 | 26,300 | 24,000 | 31,600 |
| Uni-directional 2 | 24,100 | 23,100 | 26,200 |

**Figure 8:**
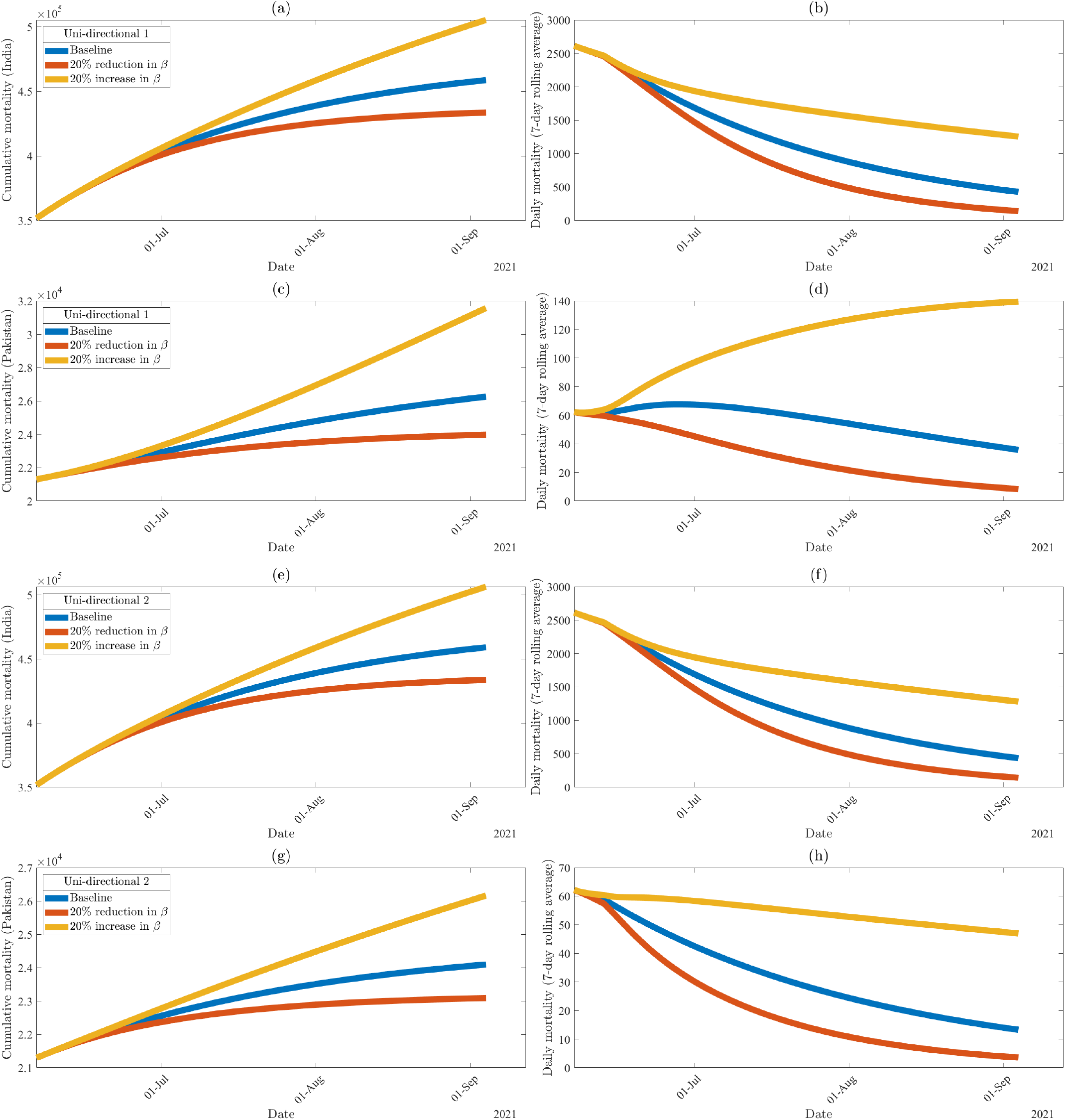
Numerical simulations of the two-patch metapopulation model (3.8) for various effectiveness levels of the implementation or relaxation of community lockdown measures under the semi-symmetric mobility scenarios listed in Table 8, showing (a-b) cumulative and seven-day rolling average daily mortality projections for India and under uni-directional 1, (c-d) cumulative and seven-day rolling average daily mortality projections for Pakistan and under uni-directional 1, (e-f) cumulative and seven-day rolling average daily mortality projections for India and under uni-directional 2, and (g-h) cumulative and seven-day rolling average daily mortality projections for Pakistan and under uni-directional 2. Initial conditions were obtained via simulation of the basic model (2.1) up to June 6, 2021. Values for fixed parameters for both countries are listed in Table 3. Values for parameters specific to each country during lockdown are listed in Table 4a (India) and Table 4b (Pakistan). Effectiveness of lockdown is measured by increasing or decreasing the estimated community contact rates (*β*_*P*_, *β*_*I*_, *β*_*A*_, *β*_*H*_) by the indicated percentage. Values obtained during the lockdown period for each country are used for other estimated parameters (*ϕ, δ*_*I*_, *δ*_*H*_).

In the uni-directional 2 mobility scenario, where residents of India do not travel, our simulations project 459, 000 and 24, 100 deaths for India and Pakistan, respectively, under the baseline lockdown scenario (with *ℛ*_0*m*_ *≈* 0.80). Furthermore, if lockdown measures are strengthened (to correspond to a 20% reduction in the community contact rate parameters), India and Pakistan could avert 25, 000 and 1000 deaths (Figures 7(e) and (g), brown curves) by September 4, 2021 (and ℛ_0*m*_ decreases to 0.64). If lockdown measures are relaxed (to correspond to the 20% increase in the contact rate parameters), our simulations show that India and Pakistan will record additional 48, 000 deaths (a 10.5% increase from baseline) and 2, 100 deaths (a 8.7% increase), respectively, by September 4, 2021. As shown in Figures 8(f) and (g), daily mortality decreases over time regardless of lockdown level (*albeit* more rapid declines are observed with baseline or stricter control measures).

Figure 9 shows new infections and daily mortality in Pakistan, as a function of time, under the four mobility scenarios where control measures are relaxed to the extent that community contact rates increase 20%. Both new cases and daily mortality in the semi-symmetric 1 (Figure 9, blue curves) and uni-directional 1 (Figure 9, gold curves) scenarios peak in September of 2021 (although a few weeks apart), with a slow decline following. In the semi-symmetric 2 scenario, where residents of both countries spend 5% of their time in the other patch, new cases in Pakistan increase rapidly increase and peak near the end of July 2021 (Figure 9(a), brown curve); daily mortality peaks in mid-August of 2021 (Figure 9(b), brown curve). In the uni-directional 2 scenario, a brief outbreak occurs almost immediately after mobility begins at the beginning of June 2021, and a small increase in daily mortality is observed later that same month with continuous declines in cases and mortality thereafter (Figure 9, purple curves).

**Figure 9:**
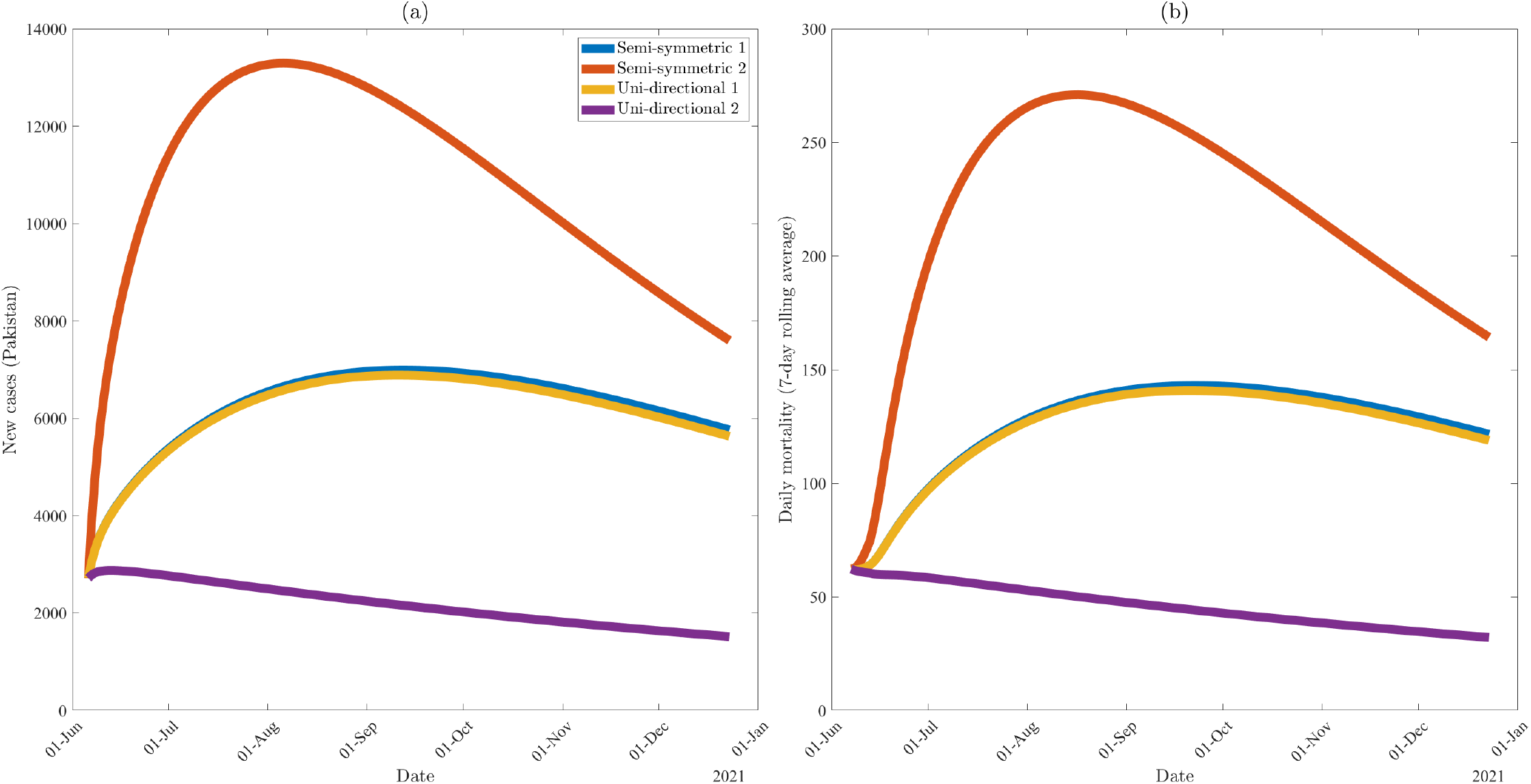
Numerical simulations of the two-patch metapopulation model (3.8) in Pakistan under various mobility scenarios listed in Table 8, where control measures are relaxed to the extent that community contact rates are increased by 20%. (a) New cases in Pakistan, (b) the seven-day rolling average of daily mortality in Pakistan. Initial conditions were obtained via simulation of the basic model (2.1) up to June 6, 2021. Values for fixed parameters for both countries are listed in Table 3. Values for parameters specific to each country during lockdown are listed in Table 4a (India) and Table 4b (Pakistan). Effectiveness of lockdown is measured by increasing the estimated community contact rates (*β*_*P*_, *β*_*I*_, *β*_*A*_, *β*_*H*_) by the 20%. Values obtained during the lockdown period for each country are used for other estimated parameters (*ϕ, δ*_*I*_, *δ*_*H*_).

The simulation results described in this section are tabulated in Tables 9 and 10. Overall, these results highlight the need to consider travel restrictions as well as the direction of travel being limited. Although projected cumulative mortality for India stays relatively the same regardless of mobility scenario, cumulative mortality projections for Pakistan may increase as much as 41% depending on the lockdown and mobility scenario (see the third column of Table 9b). Moreover, it is worth noting here that increasing the value of the time that residents of India spend in Pakistan, (*m*_*ip*_) from 0.01 to 0.05 (which indicates a shift from semi-symmetric 1 scenario to semi-symmetric 2 scenario) will result in a 17.1% increase in the projected cumulative mortality for Pakistan from the baseline. Furthermore, if the lockdown measures are relaxed, to correspond to a 20% increase in the community contact rate parameters, then Pakistan could see an increase in the additional projected cumulative deaths from 5, 400 to 10, 800 (which represents a 100% increase in cumulative deaths are projected for Pakistan, if *m*_*ip*_ increases from 0.01 to 0.05). Thus, these simulations suggest that increasing the residence-time residents of India spend in Pakistan increases the likelihood for significant outbreaks (and higher mortality) in Pakistan. However, after an initial surge of infections due to increasing residence time residence of one patch spend in the other, the disease will eventually die out in both countries regardless of the mobility scenario (since the reproduction number, ℛ_0*m*_, is less than unity in each mobility scenario). This result is consistent with the results reported in Section 2.2 for the single-patch model (2.1).

**Table 10:**
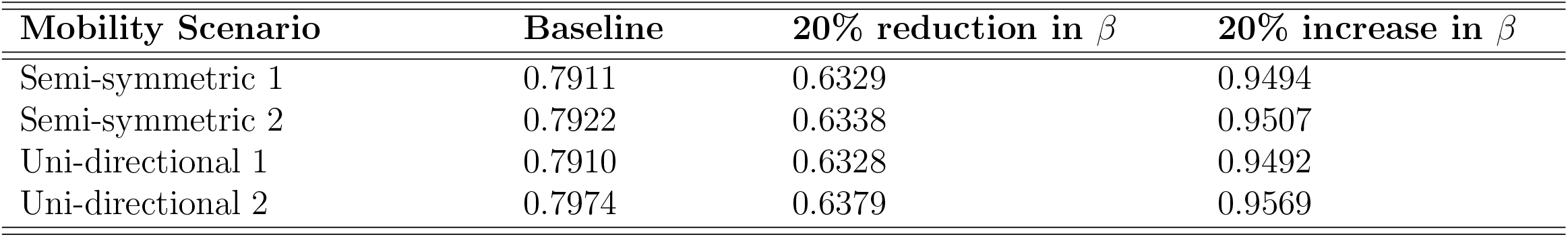
Reproduction numbers (*ℛ*_0*m*_) for the metapopulation model (3.8) under different mobility and lockdown effectiveness scenarios. Residence-time proportions for each human mobility scenario are listed in Table 8. The reproduction number is computed using the next generation operator matrices given in Appendix C. Values for fixed parameters are listed in Table 3. Values for parameters estimated during lockdown are listed in Table 4a (for India) and Table 4b (for Pakistan). Effectiveness of lockdown is measured by increasing or decreasing the estimated community contact rates (*β*_*P*_, *β*_*I*_, *β*_*A*_, *β*_*H*_) by the indicated percentage. Values estimated during the lockdown period for each country are used for other estimated parameters (*ϕ, δ*_*I*_, *δ*_*H*_).

Table 11 summarizes the projected time-to-elimination of the COVID-19 pandemic in India (Table 11a) and Pakistan (Table 11b) under the aforementioned mobility scenarios, for various effectiveness levels of mitigation measures. Time-to-elimination is measured by the date where new infections are zero asymptotically. This table shows, under the baseline level of lockdown, that the pandemic can be ended in India by October to November 2022 under all four mobility scenarios. Similarly, the model projects the end of the pandemic in Pakistan by May to July of 2022 under the baseline lockdown level for the four mobility scenarios. However, if mitigation measures are strengthened to a level that reduces the baseline levels of the community contact rate parameters by 20%, such elimination can be attained for India by March of 2022 and Pakistan by December 2021 to January 2022. If, on the other hand, lockdown measures are relaxed to the extent that the community contact rate parameters are increased by 20%, the projected timing to pandemic elimination for India extends to December 2025 to March 2026. For Pakistan, this timing extends to April to June of 2025, but may continue as long as February 2026 if measures are relaxed in the second uni-directional scenario. Although the disease is projected to persist for a longer time period in the second uni-directional scenario, Pakistan is projected to record higher cumulative mortality in the other scenarios when *m*_*ip*_ *>* 0 (i.e., residents of India are able to travel to Pakistan). It should be emphasized that, although the time-to-elimination is on a scale of three to five years, from the current time (summer 2021), the two countries will only be experiencing mild outbreaks of the pandemic during this time period. This is because, for each of the four mobility scenarios (semi-symmetric 1 and 2 and uni-directional 1 and 2) and three effectiveness levels of lockdown measures (baseline, 20% reduction in *β* and 20% increase in *β*), the reproduction number of the model is always less than unity (see Table 11). Furthermore, increased accessibility and uptake of vaccines may shorten the time-to-elimination.

**Table 11:** Estimated time-to-elimination of COVID-19 in India and Pakistan using the metapopulation model (3.8) under various lockdown and mobility scenarios. Time to elimination is measured as the time when new infections are zero asymptotically. Mobility scenarios are described in Table 8.

| Mobility Scenario | Baseline ($\mathcal{R}_{0m}$) | 20% reduction in $\beta$ ($\mathcal{R}_{0m}$) | 20% increase in $\beta$ ($\mathcal{R}_{0m}$) |
| --- | --- | --- | --- |
| Semi-symmetric 1 | Nov. 2022 (0.79) | March 2022 (0.63) | March 2026 (0.95) |
| Semi-symmetric 2 | Nov. 2022 (0.79) | March 2022 (0.63) | March 2026 (0.95) |
| Uni-directional 1 | Nov. 2022 (0.79) | March 2022 (0.63) | March 2026 (0.95) |
| Uni-directional 2 | Oct. 2022 (0.80) | March 2022 (0.64) | Dec. 2025 (0.96) |

| Mobility Scenario | Baseline ($\mathcal{R}_{0m}$) | 20% reduction in $\beta$ ($\mathcal{R}_{0m}$) | 20% increase in $\beta$ ($\mathcal{R}_{0m}$) |
| --- | --- | --- | --- |
| Semi-symmetric 1 | July 2022 (0.79) | Jan. 2022 (0.63) | June 2025 (0.95) |
| Semi-symmetric 2 | July 2022 (0.79) | Jan. 2022 (0.63) | April 2025 (0.95) |
| Uni-directional 1 | July 2022 (0.79) | Jan. 2022 (0.63) | June 2025 (0.95) |
| Uni-directional 2 | May 2022 (0.80) | Dec. 2021 (0.64) | Feb. 2026 (0.96) |

**(b)** Estimated time-to-elimination of COVID-19 in Pakistan using the metapopulation model (3.8) under various lockdown and mobility scenarios.

## Discussion

India has experienced a devastating second wave of the COVID-19 pandemic during the spring of 2021. This wave, which thoroughly overwhelmed India’s healthcare system (in addition to making India the latest global epicenter of the COVID-19 pandemic) [36, 37], is associated with the emergence of a deadly and easily-transmissible variant of the SARS-CoV-2 virus (known as the Delta variant) [39]. The objective of this project was to assess the potential impact of the devastating COVID-19 pandemic in India on the dynamics of the disease in its neighboring countries. Specifically, we sought to use mathematical modeling approaches, together with statistical data analytics, to analyse the impact of back-and-forth mobility between India and its most populous neighbor, Pakistan.

We first designed a model for the transmission dynamics of the COVID-19 pandemic in a population. The *basic model* was formulated based on stratifying the total population into a number of compartments, based on disease status. Some notable features of the basic model, which takes the form of an epidemic Kermack-McKendrick-type deterministic system of nonlinear differential equations, include allowing for disease transmission by pre-symptomatically- and asymptotically-infectious individuals (which are known to be the main drivers of the COVID-19 pandemic globally [18, 40, 65–67]. The basic model has a continuum of disease-free equilibria, which was shown to be globally-asymptotically stable whenever a certain epidemiological threshold, known as the *basic reproduction number* (denoted by ℛ_0_) is less than one. The epidemiological implication of this result is that COVID-19 outbreaks will rapidly die out in each of the two nations if the respective reproduction number can be brought to (and maintained at) a value less than one. The basic model was parameterized using cumulative COVID-19 mortality data for both India and Pakistan.

Numerical simulations of the basic model showed that relaxing the lockdown and mitigation measures in India and Pakistan, from their baseline levels, could trigger another wave of the pandemic in either country if the level of lifting or relaxation of the control measures is moderate to high (e.g., if control measures are relaxed to the extent that the baseline levels of the associated community contact rate parameters are increased by 40%). That is, relaxing current level of control measures (at moderate or high level) could cause a third and fourth wave of the pandemic in India and Pakistan, respectively. This result is consistent with that reported in [18], where the level of lifting of lockdown measures in the United States was shown to determine the size of the predicted third wave of the pandemic in the fall of 2020. Furthermore, our study estimates that the pandemic could be eliminated in India and Pakistan by July and February of 2022, respectively, if current baseline levels of control and mitigation measures are maintained.

The basic model was extended to account for the impact of the back-and-forth mobility between the two countries. The resulting two-patch metapopulation model, which considers each of the two countries as a separate patch, adopts a Lagrangian mobility pattern between the two nations (i.e., we considered movement between the nations to be of limited, not permanent, duration). Our study showed that the projected cumulative mortality for India (by September 4, 2021) remain fairly constant regardless of which of the four mobility scenarios (semi-symmetric 1 and 2 and uni-directional 1 and 2) is used in the simulations. In particular, the projected cumulative mortality for India for the baseline lockdown scenario is approximately 459,000 (this number reduces to approximately 434,000 if control measures are strengthened so that the community contact rate parameters are decreased by 20% from their baseline values). For Pakistan, the choice of mobility scenario plays an important role in determining the size of the projected cumulative mortality. For instance, our study showed a projected cumulative mortality for Pakistan (by September 4, 2021), under the baseline level of lockdown, to range from 24,000 to about 31,000 (with the lowest projected mortality achieved under the uni-directional 2 scenario, and the highest recorded under the semi-symmetric 2 scenario). Thus, for the baseline scenario, Pakistan could record more COVID-19 deaths if residents of India spend 5% of their residence-time in Pakistan and residents of Pakistan spend 5% of their residence-time in India (i.e., the semi-symmetric 2 scenario). Specifically, under this mobility scenario, Pakistan could record up to 41, 000 projected cumulative deaths by early September 2021 if control measures are relaxed to the extent that the associated community contact rate parameters are increased by 20% from their baseline values.

In summary, our study shows that the prospect of the effective control or elimination of the COVID-19 pandemic in each of the two countries, using existing control resources (based primarily on the use of non-pharmaceutical interventions), is very promising (if the interventions are maintained at their current baseline levels of coverage and efficacy). Specifically, our simulations suggest that, if control measures are maintained at their current baseline levels (and based on the mobility scenarios considered), the COVID-19 pandemic could be eliminated in India by October to November of 2022. Similarly, Pakistan could eliminate the pandemic by May to July of 2022. It should be recalled that the projected time to elimination for India and Pakistan, using the basic model (with no mobility between the two patches), were July 2022 and February 2022, respectively. Thus, the epidemiological consequence of the back-and-forth mobility between India and Pakistan (based on the mobility scenarios considered in our study) is that the time-to-elimination of the COVID-19 pandemic in India and Pakistan could be delayed by three to five months. Furthermore, the cumulative COVID-19 mortality in Pakistan increases with increasing time residents of India spend in Pakistan (*m*_*ip*_). Relaxation of control measures, combined with increasing values of *m*_*ip*_, would trigger a fourth pandemic wave in Pakistan (that is expected to peak in mid-August to mid-September of 2021).

Some of the limitations of the two models considered in this study include not explicitly accounting for the impact of some control interventions, notably vaccination. Our justification for not including vaccination is that, during the beginning of the devastating wave in India (Spring 2021), the vaccination coverage in both India and Pakistan were quite low [68]. Furthermore, the models do not incorporate some heterogeneities, such as age and risk structure, that may also be relevant to the dynamics of the disease in the two nations.

## Data Availability

The data used in this study is publicly available

## Acknowledgments

One of the authors (ABG) acknowledge the support, in part, of the Simons Foundation (Award #585022) and the National Science Foundation (Grant Number: DMS-2052363). Another author (SS) acknowledges the support of the Fulbright Scholarship.

## Appendix A Proof of Theorem 2.1

*Proof*. Consider the second equation of the basic model (2.1), *Ė* = *λS − σ*_1_*E*, which can be re-written in terms of the inequality *Ė ≥ −σ*_1_*E*, with solution *E*(*t*) *≥ E*(0) exp(*−σ*_1_*t*) *≥* 0. Similarly, the following inequalities can be established for the solutions of the next three state variables of the basic model:

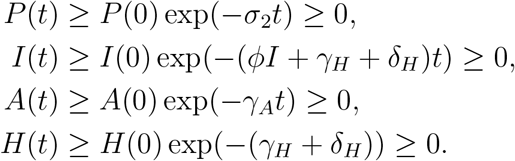

It follows from the above inequalities that, *P* (*t*), *I*(*t*), *A*(*t*), and *H*(*t*) are non-negative for all time *t >* 0; thus it can be seen that the following inequality holds for *S*(*t*) as well:

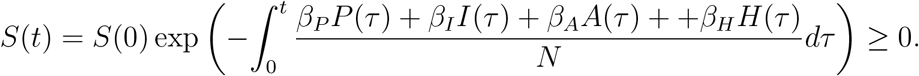

Noting that the number of recovered individuals (*R*(*t*)) is increasing (since 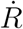 depends on *I, A*, and *H*, all of which are shown to be non-negative), then *R* is also non-negative for all *t >* 0). Thus, we have shown that the state variables (*S*(*t*), *E*(*t*), *P* (*t*), *I*(*t*), *A*(*t*), *H*(*t*), *R*(*t*)), of the basic model (2.1), are non-negative for all time *t >* 0.

Furthermore, the solutions of the basic model are bounded since *S*(*t*) + *E*(*t*) + *P* (*t*) + *I*(*t*) + *A*(*t*) + *H*(*t*) + *R*(*t*) ≤*N* (0) for all time *t >* 0. Hence, the model (2.1) is mathematically- and epidemiologically well-posed in the feasible region Ω [17].

## Appendix B Proof of Theorem 2.3

*Proof*. Consider the basic model (2.1). Further, consider the following linear Lyapunov function:

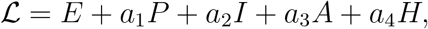

where 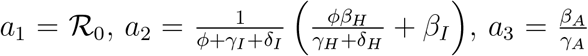, and 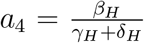. The Lyapunov derivative is given by:

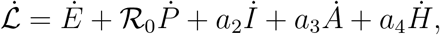

which can be simplified to

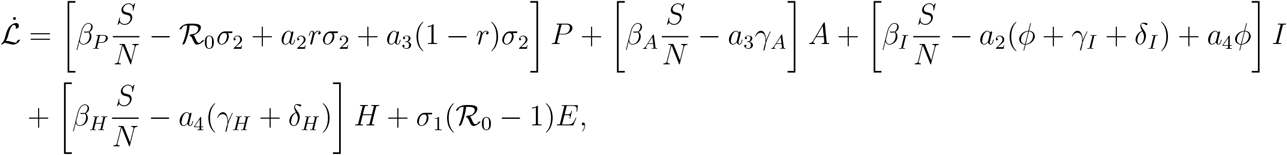

so that (noting that *S*(*t*) *≤ N* (*t*) for all *t* in Ω),

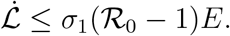

Hence, 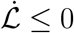 if *ℛ*_0_ *≤* 1, and 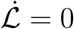 if and only if *E*(*t*) = 0. Substituting *E*(*t*) = 0 into the equations of the basic model (2.1) show that (*S*(*t*), *E*(*t*), *P* (*t*), *I*(*t*), *A*(*t*), *H*(*t*), *R*(*t*)) *→* (*N* (0)*−R*^*∗*^, 0, 0, 0, 0, 0, *R*^*∗*^) as *t → ∞*. Furthermore, it can be shown that the largest compact invariant set in 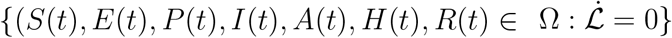 is the continuum of disease-free equilibria (*ξ*_0_). Hence, it follows, by LaSalle’s Invariance Principle [49], that the continuum of disease-free equilibria (*ξ*_0_) of the basic model (2.1) is globally-asymptotically stable in Ω whenever *ℛ*_0_ *≤* 1.

## Appendix C Computation of Reproduction Number

Here, too, the next-generation operator method will be used to compute the reproduction number for the metapopulation model (3.8), denoted by ℛ_0*m*_. It can be seen that the associated matrix of new infection terms of the metapopulation model (denoted by *F*) is given by:

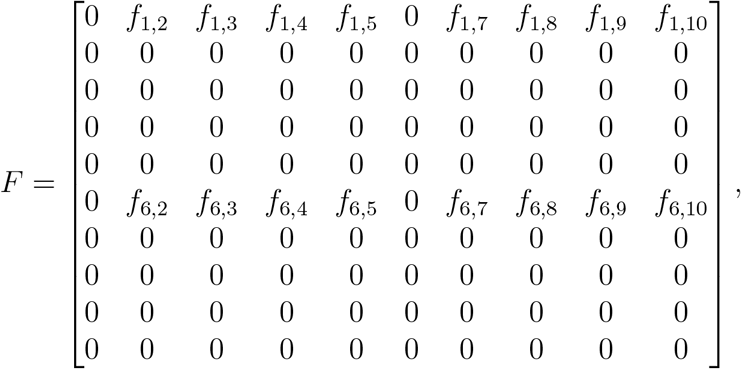

where,

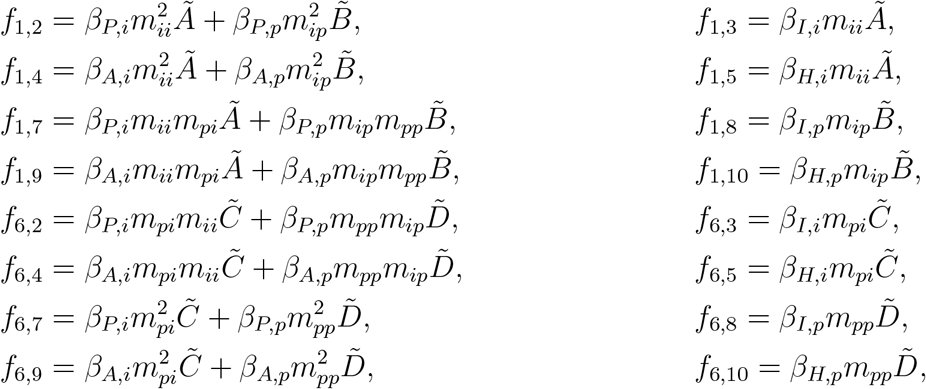

with,

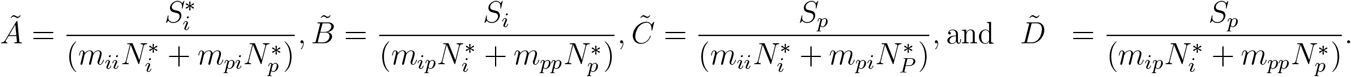

Similarly, the associated matrix of linear transition terms in the infected compartments of the metapopulation model (denoted by *V*) is given by:

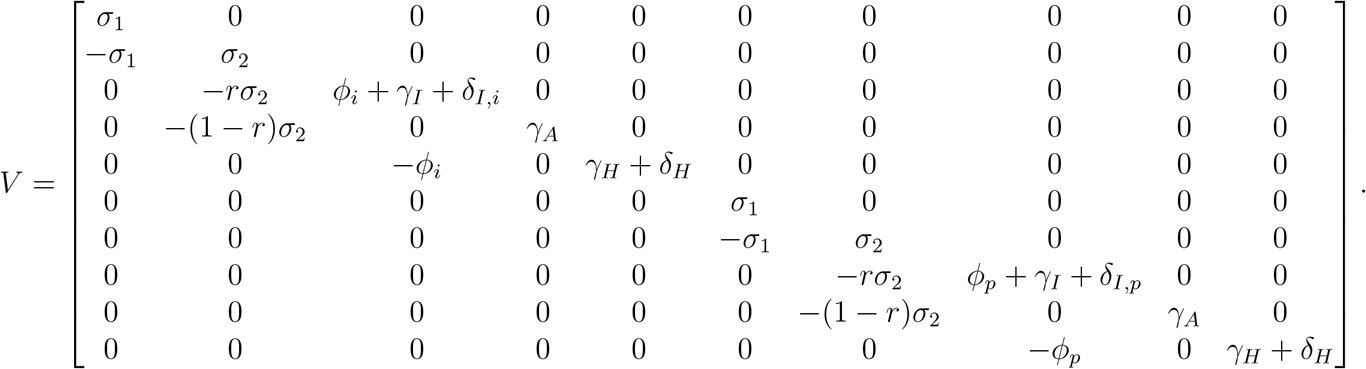

Thus, it follows from [48] that the reproduction number of the metapopulation model (3.8) is given by

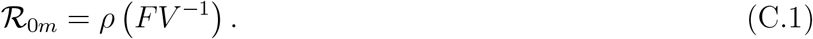

It should be stated that, although the reproduction number (ℛ_0*m*_) for the metapopulation model, given by (C.1), could not be readily expressed in closed form (owing to the large size of the next generation matrix *FV* ^*−*1^), we compute it numerically (by directly substituting the values of the parameters involved in each entry of the two matrices) for the various scenarios discussed in Section 3.4.2.

